# Portable Multi-focal Visual Evoked Potential Diagnostics for Multiple Sclerosis/Optic Neuritis patients

**DOI:** 10.1101/2023.12.26.23300405

**Authors:** S. Mohammad Ali Banijamali, Craig Versek, Kristen Babinski, Sagar Kamarthi, Deborah Green-LaRoche, Srinivas Sridhar

**Affiliations:** Department of Mechanical and Industrial Engineering, Northeastern University, Boston, MA, USA; NeuroFieldz Inc, Newton, MA, USA; Department of Neurology, Tufts University School of Medicine, Tufts Medical Center, Boston, MA, USA; Department of Physics, Department of Bioengineering and Department of Chemical Engineering, Northeastern University, Boston, MA 02115

**Keywords:** Multi-Focal Visual Evoked Potential (mfVEP), Full-Field Visual Evoked Potential (ffVEP), Multiple Sclerosis (MS), Optic Neuritis, Machine Learning, Signal Processing, Portable Diagnostics, Support Vector Machine (SVM)

## Abstract

**Purpose:** Multiple Sclerosis (MS) is a neuro-inflammatory disease of the Central Nervous System (CNS) in which the body’s immune system attacks and destroys myelin sheath that protects nerve fibers and causes disruption in axonal signal transmission. Demyelinating Optic Neuritis (ON) is often a manifestation of MS and involves inflammation of the optic nerve. ON can cause vision loss, pain and discomfort in the eyes, and difficulties in color perception.

In this study, we developed NeuroVEP, a portable, wireless diagnostic system that delivers visual stimuli through a smartphone in a headset and measures evoked potentials at the visual cortex from near the O1, Oz, O2, O9 and O10 locations on the scalp (extended 10-20 system) using custom electroencephalography (EEG) electrodes.

**Methods:** Each test session is constituted by a short 2.5-minute full-field visual evoked potentials (ffVEP) test, followed by a 12.5-minute multifocal VEP (mfVEP) test. The ffVEP test evaluates the integrity of the visual pathway by analyzing the P1 (also known as P100) component of responses from each eye, while the mfVEP test evaluates 36 individual regions of the visual field for abnormalities. Extensive signal processing, feature extraction methods, and machine learning algorithms were explored for analyzing the mfVEP responses. The results of the ffVEP test for patients were evaluated against normative data collected from a group of subjects with normal vision. Custom visual stimuli with simulated defects were used to validate the mfVEP results which yielded 91% accuracy of classification.

**Results:** 20 subjects, 10 controls and 10 with MS and/or ON were tested with the NeuroVEP device and a standard-of-care (SOC) VEP testing device which delivers only ffVEP stimuli. In 91% of the cases, the ffVEP results agreed between NeuroVEP and SOC device. Where available, the NeuroVEP mfVEP results were in good agreement with Humphrey Automated Perimetry visual field analysis. The lesion locations deduced from the mfVEP data were consistent with Magnetic Resonance Imaging (MRI) and Optical Coherence Tomography (OCT) findings.

**Conclusion:** This pilot study indicates that NeuroVEP has the potential to be a reliable, portable, and objective diagnostic device for electrophysiology and visual field analysis for neuro-visual disorders.

## Introduction

Multiple Sclerosis (MS) is a neuroinflammatory disease that damages the myelin sheath, nerve cell bodies, and axons. The resulting lesions may be found throughout the spinal cord and the brain. Optic Neuritis (ON) is often a manifestation of MS and involves inflammation of the optic nerve, causing partial or complete loss of vision usually in one eye. About 1 in 5 MS patients experience ON as their initial symptom, and 50 percent of people with MS experience ON during the course of their disease [1]. More than 2.3 million people are thought to have MS worldwide, and the disease has impacted more than 400,000 people in the USA [2].

Visual evoked potentials (VEPs) are electrophysiological signals in response to visual stimuli that are retrieved from the electroencephalographic activity in the visual cortex and are recorded using scalp electrodes. Responses from a full-field VEP (ffVEP) test, which appear significantly delayed compared to normal, have long been known to be a biomarker diagnosing MS [3]. The latency of the P1 component of ffVEP responses provides reliable and objective information about the integrity of the visual system and proof of demyelination in MS patients, but is limited in terms of providing spatially localized information about the visual field. The multi-focal visual evoked potentials (mfVEP) technique enables the simultaneous recording of VEP responses from a large number of visual field regions in a time much shorter than sequential methods, allowing for the identification of spatially localized retinal damage as well as the lateralization of optic nerve and cortical dysfunction with the use of multi-channel recording montages [4]. This technique has proven applications in the assessment of visual field disorders and the diagnosis of diseases like optic neuritis/multiple sclerosis and glaucoma [5–7]. ffVEP response quality can provide a useful guide to interpreting the more complex mfVEP signals; therefore, we hypothesized that combining these two methods would yield a more powerful diagnostic tool than either alone.

Utilizing a custom mobile virtual reality (VR) headset and in-house developed neuroelectric sensing hardware, we have developed a portable wireless system, called NeuroVEP, that enables the combined ffVEP and mfVEP tests. Using a mobile phone display, stimuli are presented to each eye separately while the other is held in darkness. An array of eight proprietary hydrogel electrodes senses the brain’s responses in the vicinity of O1, Oz, O2, O9 and O10 scalp locations (extended 10-20 system). Recording from multiple locations allows for the lateralization of ffVEP responses coming from the left and right hemispheres and helps with improving the signal-to-noise ratio (SNR) for mfVEP results. Using the Scientific Python and Scikit Learn platform [8, 9], we developed an automated signal processing, statistical analysis, and machine-learning framework, which handles our recorded EEG signals and stimulus timing events for the combined ffVEP and mfVEP testing paradigms.

The device was tested on a group of subjects with MS and/or ON and a control group of normally sighted subjects. The ffVEP results were compared to a standard of care (SOC) device for ffVEP testing, Natus Nicolet, used by our clinical partners. Unlike the NeuroVEP which is portable, the Natus system is cart-based; furthermore, the SOC’s electrodes are slightly invasive, requiring needles to be inserted into the subject’s scalp.

## Materials and Methods

### Subject Pool

This study was approved by the Northeastern University and Tufts Medical Center Institutional Review Boards (IRB Study # 13395, Protocol title: Objective Portable Diagnostics of Neurological Disorders using Visual Evoked Potentials) and was performed in accordance with the Declaration of Helsinki. All subjects were either referred by their clinical neurologist at the Department of Neurology at Tufts Medical Center or were self-enrolled after seeing an advertisement posted around Tufts Medical Center. Subjects were required to be between 18 and 80 years of age and signed a written informed consent document after the study was explained and all of their questions were answered. Gender was not used as a condition for selection. 10 healthy subjects along with 10 subjects with MS, ON or both conditions were tested.

Because of the virtual reality (VR) capabilities of our display system, we required subjects to fill out a simulator sickness questionnaire; however, the static nature of our stimuli was not expected to induce motion sickness symptoms, and this was borne out in the results (no discomfort was reported in the subject feedback). Three of the authors served as control subjects (SMAB, CV, and SS), who have extensive experience participating in EEG and/or psychophysical vision testing paradigms.

The ffVEP and mfVEP experiments were tested in 10 (×2 eyes) Normal subjects and 10 (×2) subjects with MS, ON or both. A detailed overview of subject characteristics is reported in the “Results and Discussion” section (see **Table 5**).

### Hardware

We have developed a wireless headset based on mobile virtual reality technology, where a smartphone with an OLED screen displays stimuli while an array of eight neuroelectric sensors attached to the rear of the headset capture the subject’s responses from the visual cortex. We are using an updated prototype similar to the one that was developed in our lab for the diagnosis of age-related macular degeneration (AMD) [10, 11]; the modifications include: a more ergonomic fit, decreased weight, and better ventilation for the headset; the incorporation of a commercial eye-tracking device (Pupil Labs VR/AR add-on [12]); and a new generation of our custom hydrogel electrodes.

### Headmount

The headset is primarily made of parts that were 3D printed using a Formlabs Form 3 stereolithographic system. The design mounts a smartphone to the front using a switchable adapter plate and positions sensors on the back with an adjustable tension mechanism to allow consistent electrode contacts at precise scalp areas. A combination of rigid and flexible materials have been used to ensure a comfortable and light-tight fit on the head. The interior temperature and the headset’s overall weight were two other crucial factors that received particular attention. The headset also features a phototransistor sensor for accurate timing of stimulus onset and reversal and eye-tracker cameras to ensure the subjects’ compliance to test protocols.

### Neuroelectric Sensing

The NeuroVEP system makes use of custom reusable hydrogel electrodes that are both comfortable and convenient to set up. The tip of the soft yet mechanically robust hydrogel electrode (see **Figure 1.B.**) is textured in order to hold a small layer of liquid electrolyte, which penetrates through scalp hair, providing a stable and low impedance contact. The tip diameter is between 6-7mm, smaller than standard 10mm EEG cup electrodes. We have previously established [13] that contact impedances (at 30Hz) below 150 kOhms give adequate low noise levels, where values of 50-100 kOhm after touch-ups with additional electrolyte are typical.

**Figure 1.**
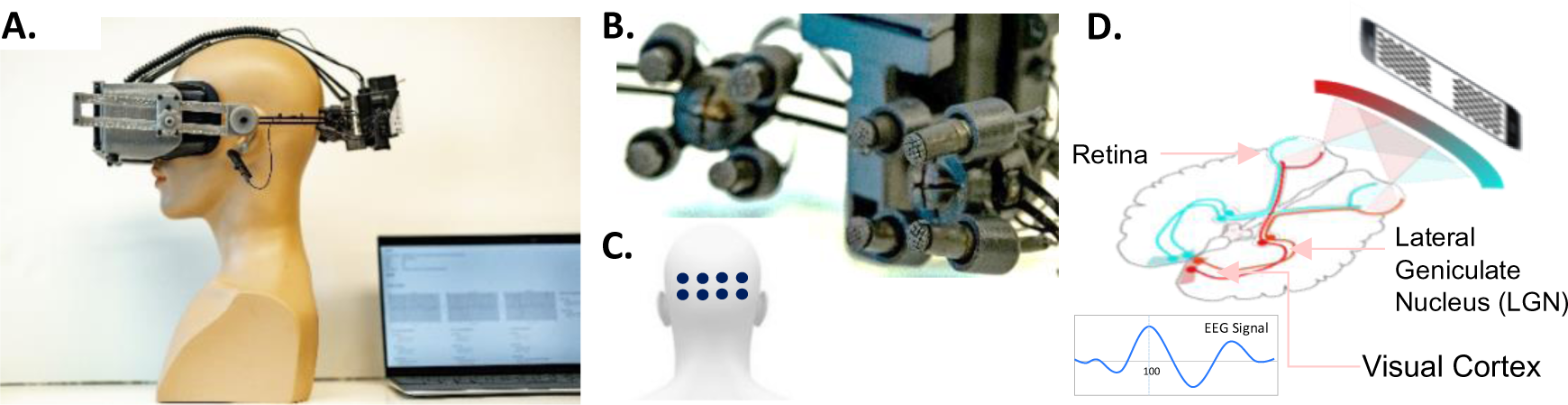
Portable wireless NeuroVEP system. **A.** NeuroVEP device prototype integrating NeuroVEP sensor with visual stimulus headset, **B.** Closeup of NeuroVEP EFEG sensor arrays, **C.** Location of electrodes on the scalp over the visual cortex, **D.** Dichoptic stimulus and typical neuroelectric response, Visual pathway. Image in part D: Derived from (https://commons.wikimedia.org/wiki/File:Human_visual_pathway.svg)

### Visual Stimuli

There are two sections to the test: a short 2.5-minute ffVEP test is followed by a longer 12.5-minute mfVEP test. The two tests are separated by a 30-second break period, and this makes the total length of the test 15.5 minutes. For the ffVEP test, a total of 90 trials are recorded for each eye, with intervals of 0.5 s plus a random 0 to 0.1 s between stimulations; the trials are broken into 6 segments of 30 repetitions, switching eyes every new segment where there is a discarded onset/offset period of 2 seconds. The mfVEP test has a total of 16384 overlapping trials at a rate of 60 frames per second; the sequence is divided into 16 segments of 1024 trials, switching eyes every new segment with a similar onset/offset period at the start.

The Google Pixel 2 XL smartphone’s OLED screen combined with optics that resemble the Google VR Daydream View (first version, 2016) was used for displaying the stimulus. Given the light-tightness of our headset and minimal reflected light in the viewer, the OLED screen on the phone offers an effectively infinite contrast ratio. The stimuli patterns were generated as high-resolution pixel maps (2048×2048) using custom Python programs, then were loaded into a custom app using the Google VR Android SDK with OpenGLES2 texture rendering.

The mfVEP stimulus is similar to a dartboard that extends to 22.25° eccentricity and is broken into 36 visual field subregions [14] (**Figure 2.B.**). The sector sizes are scaled based on cortical magnification factors to produce relatively similar cortical stimulation. Sectors are reversed (or not) simultaneously according to 36 pseudorandom uncorrelated sequences at 60 frames per second; individual responses are extracted using a correlation between each sector sequence’s time markers and the continuous EEG recording, known as the m-sequence technique [15]. The ffVEP stimulus is the same geometry dartboard, but the whole pattern reverses at the same time (**Figure 2.A.**).

**Figure 2.**
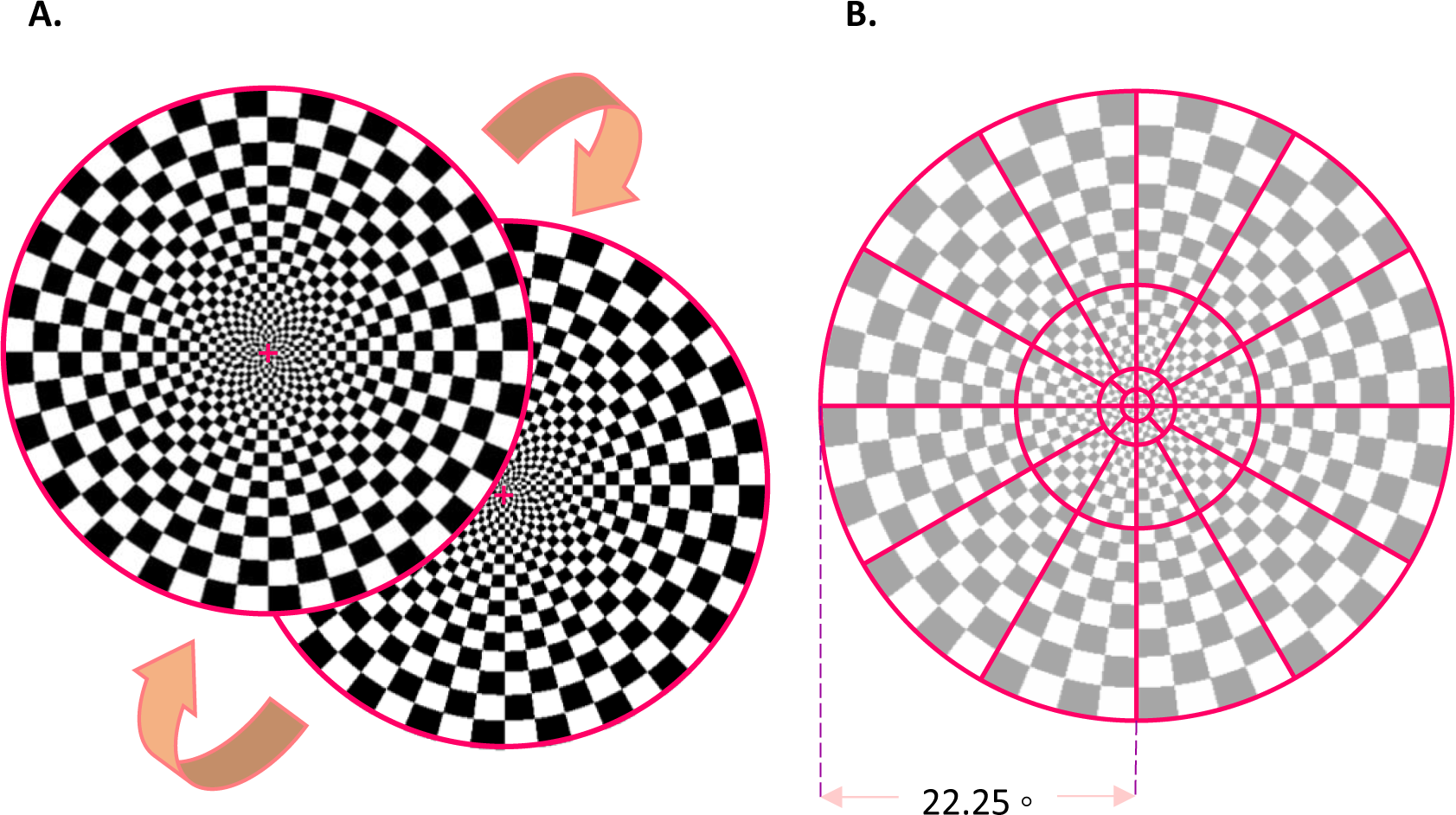
**A.** ffVEP dartboard pattern-reversal visual Stimulus, **B.** mfVEP 36 sectors visual stimulus.

### Data Analysis

The EEG recording (at 1000 samples per second) and paradigm event data are streamed wirelessly to a computer as it is acquired and are cached into a file based on the Advanced Scientific Data Format (ASDF) [16], along with subject metadata and electrode impedance measurements. On the computer, an automated data analysis framework does the work of post-processing, analysis, and classification separately on the ffVEP and mfVEP signals.

Before subsequent data processing, each channel is prefiltered using a Stationary Wavelet Transform (SWT) baseline removal with an effective cutoff frequency of 0.5 Hz; this step reduces the effect of impulse response artifacts in subsequent filtering steps [10]. For this study, where the responses will be primarily analyzed using machine learning algorithms that may be sensitive to excess noise, we made the choice to reduce the frequency content to the band of 3-13Hz (which includes P1 deflections as well as intrinsic alpha wav’es), using a Bessel high-pass IIR and an SWT low-pass filter. It is important to recognize that bandwidth reduction will inevitably create some distortion of time domain features, such as the latency of the P1 peaks; however, many other experimental aspects, such as the details of the stimuli presentation will also affect the distributions of response parameters – this is precisely why each laboratory or device system must have its own set of normative data to compare against [17].

### Full-Field Neuro-responses Signal Processing

The steps involved in the processing of the full-field responses are shown in **Figure 3**. The raw data initially undergoes the bandwidth reduction filters described above. Then, an unsupervised machine learning outlier rejection algorithm (Scikit-Learn’s “Isolation Forest” model [18]) excludes trials with outlier variances which are mainly caused by movement artifacts. Next, an alpha wave sensitive outlier rejection algorithm does a Fast Fourier Transform (FFT) on individual trials and excludes those with power predominantly within the alpha frequency band (9∼12 Hz).

**Figure 3.**
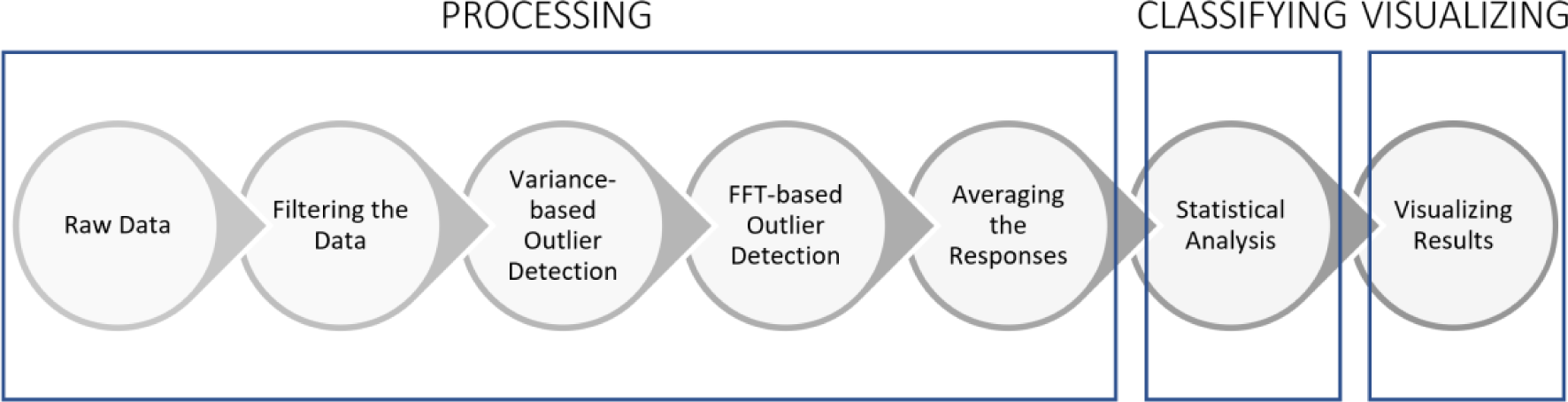
Full-Field Stimulus Data Analysis Steps.

It has been shown that alpha waves are amplified during resting, sleeping, eye closures and in our experience, when the subject is unable to see the stimulus; conversely, alpha waves are attenuated by visual attention [19–21]. Then, the selected trials recorded from each electrode are averaged across time to form 8 individual response channels. Using these 8 signals we then calculate responses from the three scalp locations: the left hemisphere is the average of electrodes 1 and 2 (O1), the right hemisphere is the average of electrodes 7 and 8 (O2) and the central signal is calculated by averaging electrodes 3, 4, 5 and 6 (Oz) (**Figure 4**).

**Figure 4.**
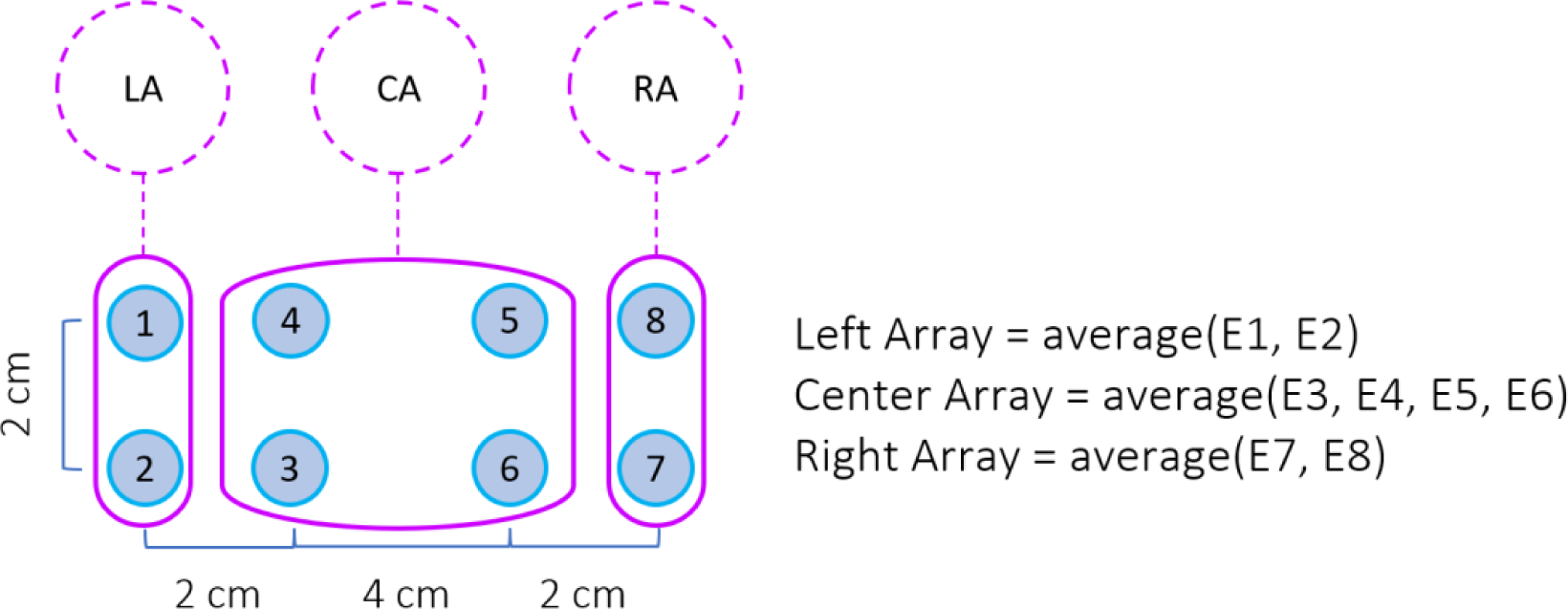
Arrangement of the electrodes and calculation of left, center and right array responses.

The first major positive component of the full-field VEP response, P1, has been shown to be delayed or absent in patients with MS or ON [3, 22, 23]. We measure the latency and amplitude of P1 components of individual eyes at three different scalp locations (O1, Oz, and O2). We also calculate the interocular and interhemispheric latency difference and amplitude ratios. Waveform and amplitude abnormalities also have been considered biomarkers for MS and ON [24].

### Full-Field Classification and Visualization

**Figure 5** shows an example of a full field report. The data are presented separately for each eye and for three separate scalp locations. Data from the left hemisphere is the average of electrodes 1 and 2 (O1), the central location is the average of electrodes 3, 4, 5, and 6 (Oz), and the data from the right hemisphere is the average of electrodes 7 and 8.

**Figure 5.**
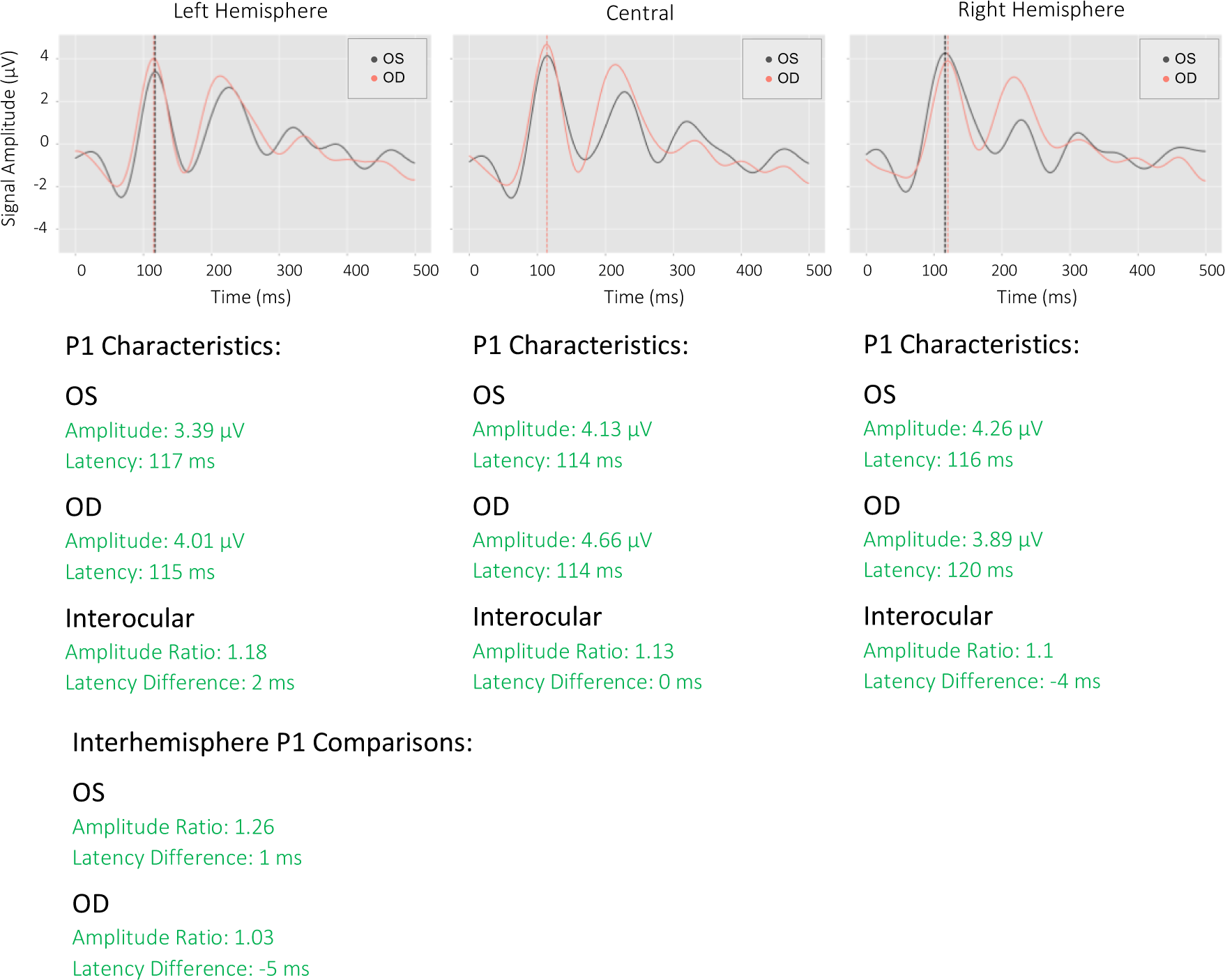
Full-field test reports of a representative normal subject from NeuroVEP Device.

6 distinct parameters have been investigated for the ffVEP signals: latency, amplitude, interocular latency difference, interocular amplitude ratio, interhemispheric latency difference, and interhemispheric amplitude ratio. Each of these parameters may indicate dysfunctions in the prechiasmatic, chiasmatic, or postchiasmatic regions of the visual field pathway.

Latency-based biomarkers are less sensitive to retinal and ocular diseases and more reliable in examining visual pathways compared to amplitude-based ones. However, in the absence of other retinal diseases, latency abnormalities generally signify demyelination whereas amplitude abnormalities are a sign of axonal loss. [19].

A monocular delayed latency suggests a dysfunction in the optic nerve on one side. Conversely, if there is a bilateral abnormality in latency, it indicates a dysfunction in the visual pathway on both sides, but determining whether it is located in pre or post -chiasmatic regions would require additional evaluation of amplitude and topographic (visual field mapping) features. [25]

Amplitude-based metrics of P1 are much more sensitive to ocular and retinal disease. Patient factors such as poor fixation, loss of focus, tearing, inattention, or drowsiness can all cause a decrease in mid-occipital amplitude. However, after ruling out these factors, a monocular amplitude abnormality suggests a dysfunction in the visual pathway of one eye before the optic chiasm. A bilateral abnormality indicates bilateral disease, but more detailed analysis of topographic features or responses to partial field stimulation is necessary to localize the specific site affected beyond the post-chiasmal region. Testing one or both eyes may reveal lateral occipital amplitude asymmetries (interhemispheric amplitude ratio abnormalities). In the absence of P1 latency or interocular amplitude abnormalities, such asymmetries are not necessarily of clinical importance. However, in the presence of the mentioned abnormalities, they may be the sole indication of dysfunction in the chiasmal or post-chiasmal visual pathway. Typically, such abnormalities are present in both eyes. If the asymmetry is unilateral, it may suggest a partial dysfunction before the optic chiasm.

To evaluate all parameters, each laboratory or device must establish its own unique normative values [17]. We tested a control group of 13 normal subjects (26 eyes) with a mean age of 33±7 years to establish classification criteria for all parameters. Based on the following criteria, each parameter is classified as normal (green), borderline (yellow), or abnormal (red), where *std* refers to its standard deviation: for all latency-based metrics, including interocular and interhemispheric latency differences, a value below “mean + 2 *std”* was marked as normal, between 2 to 3 *std* from the mean was marked as borderline, and above 3 *std* away from the mean was marked as abnormal. For all amplitude values, a natural logarithm of one plus the distribution (*ln(amp_i_+1)*) was first calculated to get a more normal distribution, then similarly, any nonexistent (or negative amplitude) P1 or values below “mean – 3 *std”* were marked as abnormal, between “mean – 3 *std”* and “mean – 2 *std”* were marked as borderline and above “mean – 2*std”* were marked as normal. This procedure has been recommended by American Clinical Neurophysiology Society Guidelines on Visual Evoked Potentials [25]. The main ffVEP statistics for the control subject pool are summarized in **Table 1**.

**Table 1.** Normative (Control group) statistics of main ffVEP parameters on 3 scalp locations. The logs of amplitudes are calculated as the natural logarithm of (1+variable).

| Stat | LA Lat | LA Lat<br>Diff | LA<br>log(Amp) | CA Lat | CA Lat<br>Diff | CA<br>log(Amp) | RA Lat | RA Lat<br>Diff | RA<br>log(Amp) |
| --- | --- | --- | --- | --- | --- | --- | --- | --- | --- |
| **Mean** | 110 | 5.08 | 1.36 | 105 | 3.31 | 1.61 | 107 | 4.77 | 1.36 |
| **Std** | 10.34 | 5.44 | 0.44 | 7.43 | 2.72 | 0.42 | 10.91 | 3.94 | 0.44 |

**LA:** Left Array, **Lat:** Latency, **Diff:** Difference, **CA:** Central Array, **Amp:** Amplitude, **RA:** Right Array

For all amplitude ratios, based on the distributions, the values were classified based on thresholds. In all cases, the ratios were calculated as the larger value to the smaller value, and then ratios falling above 2.5 were marked as abnormal, between 2 to 2.5 were marked as borderline, and less than 2 were classified as normal.

### Signal Processing of Multi-focal Neuro-visual responses

Multifocal data analysis is more complicated. The steps involved in the data processing of mfVEP data are shown in **Figure 6**. Similarly to ffVEP, the mfVEP data is filtered for bandwidth reduction (3-13Hz) and the same outlier rejection algorithm mark trials for exclusion from response averages. Next, individual sector responses are extracted using the m-sequence technique [26].

**Figure 6.**
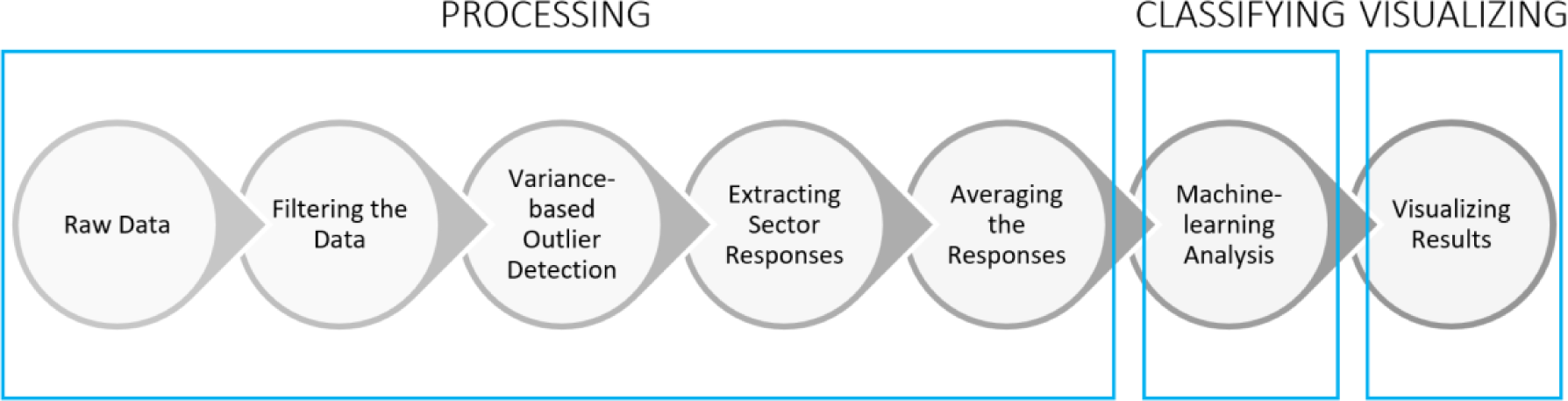
Multi-focal Stimulus Data Analysis Steps.

We take advantage of recording from 8 individual sensors to increase the signal quality. Given the variations in the folding of the brain’s visual cortex among individuals, it has been shown that recording from multiple locations and then combining the responses can significantly improve the SNR [27–29]. **Figure 7.A.** shows how derived signals D1, D2, D3, and D4 are computed from the original 8 channels. Based on our experience and recommendations from previous studies, these four combinations provided the highest SNR signals [27, 30]. **Figure 7.B.** Shows the method of calculating the SNR values. Our selection of response window and noise window is similar to what was done in the previous studies [31]. However, our calculation of the SNR and noise window criteria is slightly different. The main difference is that instead of using the average of noise windows of all sectors for each subject as the noise, we use the noise region of each individual sector of each subject for the calculation of the SNR.

**Figure 7.**
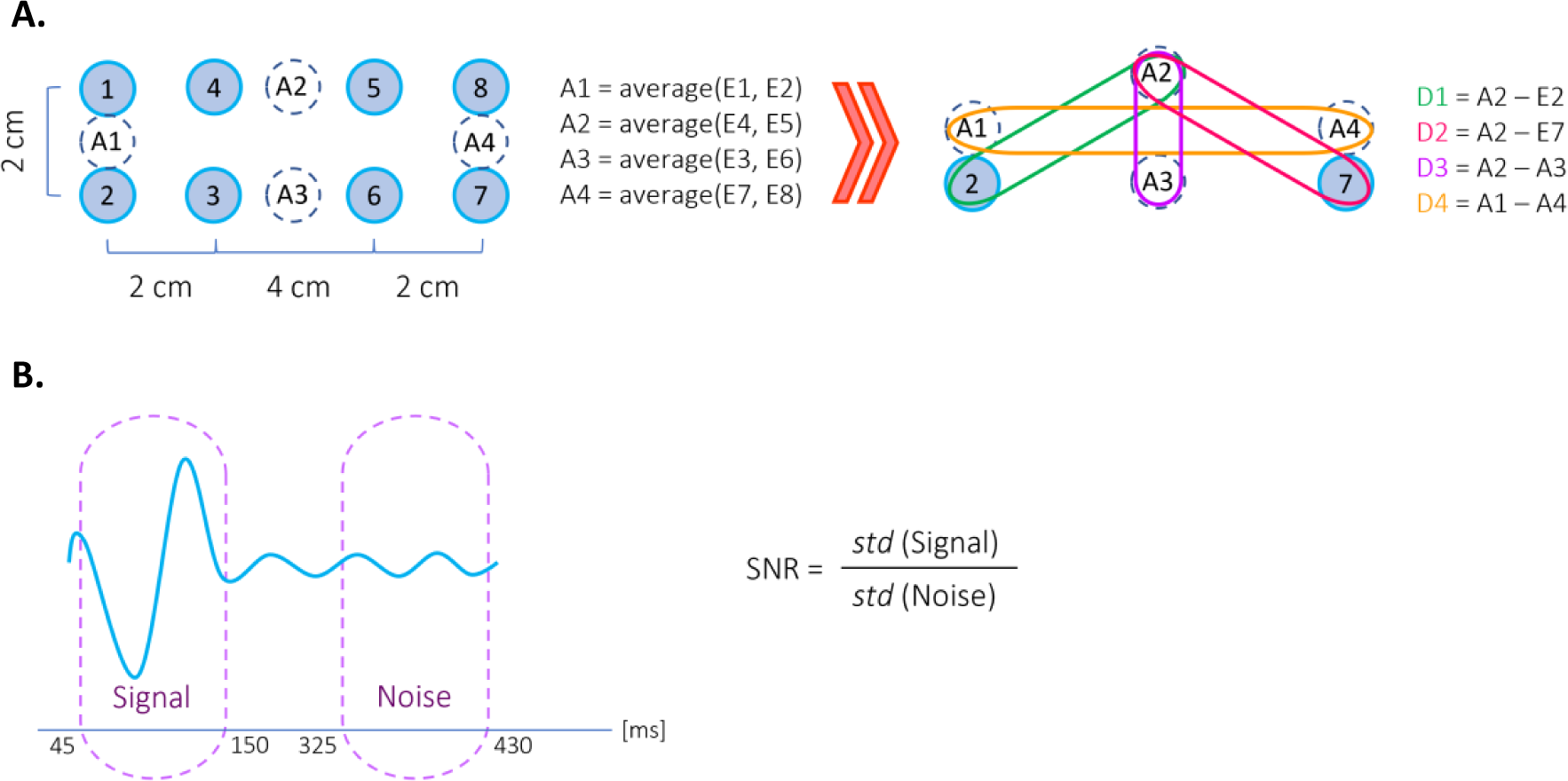
**A.** Multi-focal test electrode arrangement and calculation of derived signals. **B.** Calculation of SNR for mfVEP responses: Signal window [45:150] ms and Noise window [325:430] ms.

Finally, we compute the optimized response for each sector in the following manner: the derived signal responses are polarity matched to D2 and then are averaged together using their SNR as a weighting factor.

### Multi-Focal Data Classification

We have developed a machine learning framework for classifying and quantifying the responses. We have also created unique color maps to visualize mfVEP results.

#### Machine Learning Data Set Curation

For training the supervised Machine Learning Algorithm (MLA), we started by manually selecting and labeling the most apparent sector responses from 7 different experiments (14 eyes) as “normal” and “abnormal” - a binary classification task. The selected experiments included 6 cases of healthy subjects with artificial defects introduced to the stimulus plus 1 case of MS subjects.

The artificial defects were designed by partially or fully masking individual mfVEP sectors with black patches directly using the paradigm presentation software; we hypothesize that this masking would adequately simulate the effects of loss of vision in those regions. Responses from the masked regions or abnormal responses from the MS subject were labeled as “abnormal” and the healthy responses were labeled as the “normal” category. This initial dataset included 382 samples; however, we established that we could increase the accuracy of the machine learning algorithm by using data augmentation techniques to greatly increase the number of training samples. The training data was augmented to 65,536 (2^16^) samples using a randomized nearest-neighbor mixing and rescaling approach, where a random signal is selected and linearly interpolated with its nearest neighbor using a Gaussian random variable mixing factor centered around 0.5 with a standard deviation of 0.25, clipped to range [0,1]. We employed the KDtree algorithm [32] from the SciPy’s package for Python [9] for efficient nearest-neighbor identification.

Despite sophisticated signal processing methods that do very well in filtering artifacts and alpha waves in some individuals, it has been observed that algorithms fail to avoid false positive or false negative classifications. This issue is not unique to mfVEP – the Humphrey Visual Field test can suffer a similar problem from subject participation errors. Other researchers have tried various methods to deal with these issues for monocular mfVEP test evaluation [33, 34]. Because of the risk of over-fitting, where MLA accuracy during training is much higher than when applied to new tests, the final evaluation of the system must not use data encountered during training. To evaluate the performance of both the MLA and the mfVEP test conducted by the NeuroVEP device, we built a separate dataset based on custom mfVEP stimuli with artificial defects (shown in **Figure 8**) containing 576 samples from both eyes of 2 healthy volunteers (4 tests each, 16 tests total). In this case, the samples were not selected manually; instead, they were labeled based on whether the sector was masked or not.

**Figure 8.**
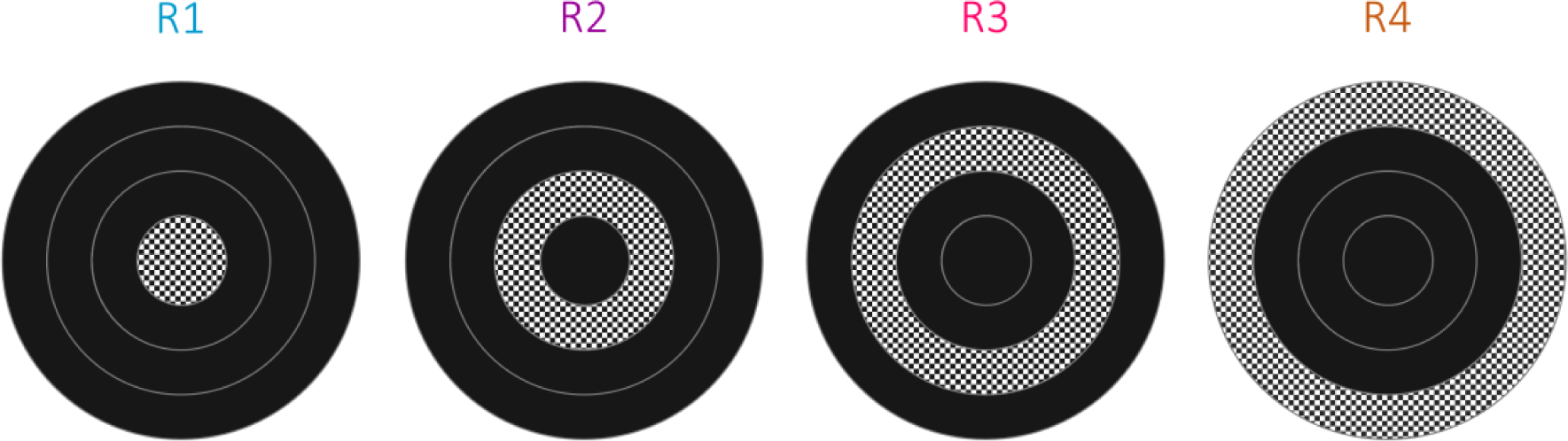
Schematic of the stimuli used for creating the “artificial defect” evaluation dataset. In each case, only sectors on one ring were unmasked (stimulating).

#### Feature Extraction

Feature extraction is a crucial step in developing a high-performance machine learning model. We started by extracting a large database of custom features which could be generally categorized into two types, Waveform-based and Frequency-based features. In the visual field regions where the subjects are unable to see (or just barely perceive) the stimulus, they usually produce responses within the frequency range of alpha waves (8∼13 Hz); this effect can be seen in the waveforms – for “abnormal” responses (which are often indistinguishable from noise), the signal and noise windows that are used for SNR calculation look very similar to each other and contain higher amplitude alpha band content, than the same windows of “normal” responses.

**Figure 9** shows 2D histograms of the two classes of responses, where the overlaid curves are binned to produce a color-scaled counts dimension.

**Figure 9.**
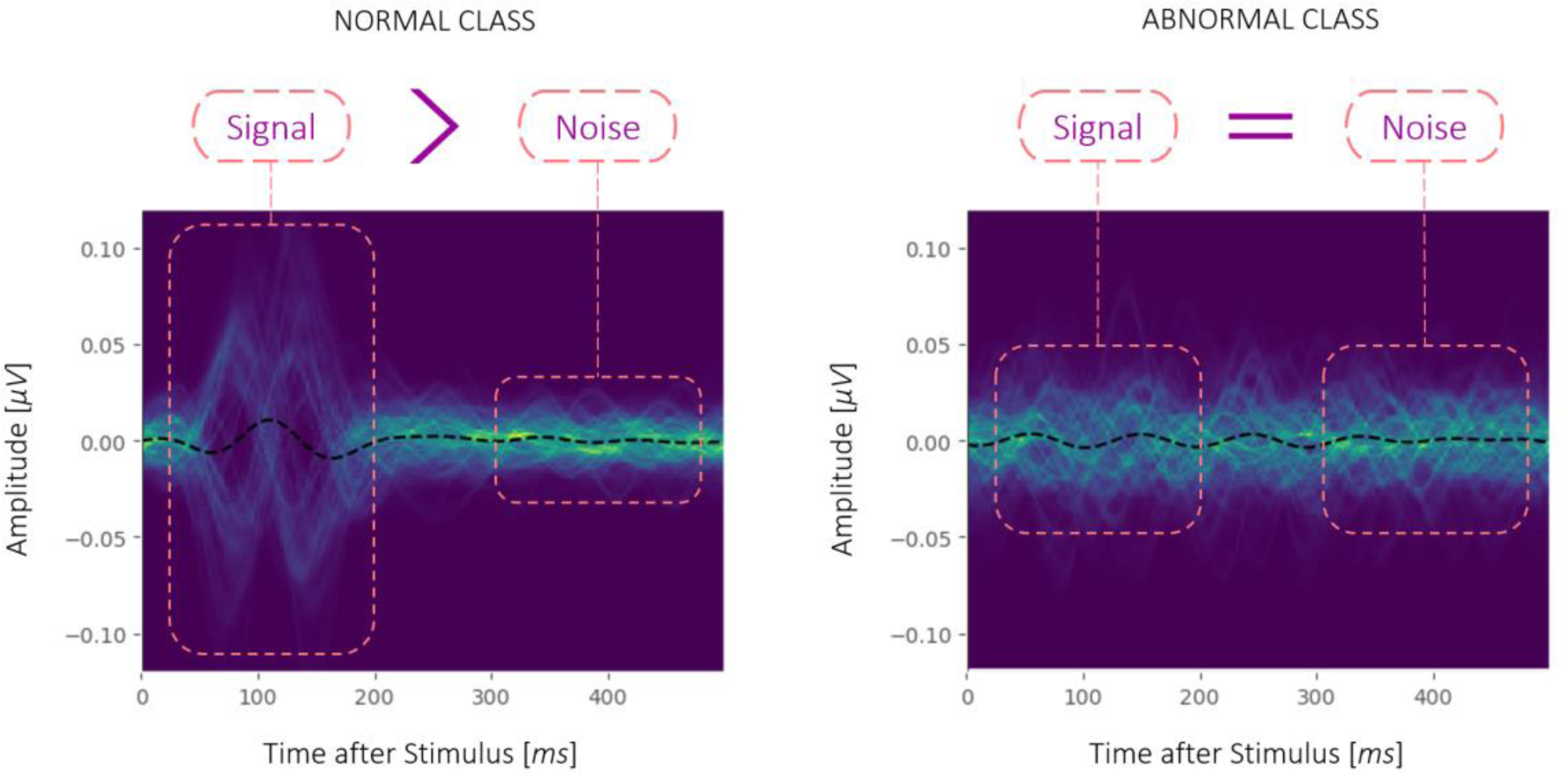
Overlay of training data **A.** Normal class vs. **B.** Abnormal class.

Based on these considerations we kept 12 custom features which could effectively discriminate between the two classes of our machine learning problem. Later, we found that we could further improve the accuracy and performance of our model by introducing 7 more features from the well-known Catch-22 feature set [35]. These features belong to the following categories: Linear autocorrelation (CO_f1ecac and CO_FirstMin_ac), successive differences (MD_hrv_classic_pnn40, SB_BinaryStats_diff_longstretch0, and SB_MotifThree_quantile_hh) and simple temporal statistics (DN_OutlierInclude_p_001_mdrmd and DN_OutlierInclude_n_001_mdrmd). Therefore, for our final machine learning model, we used 19 features, listed in **Table 2**.

**Table 2.**
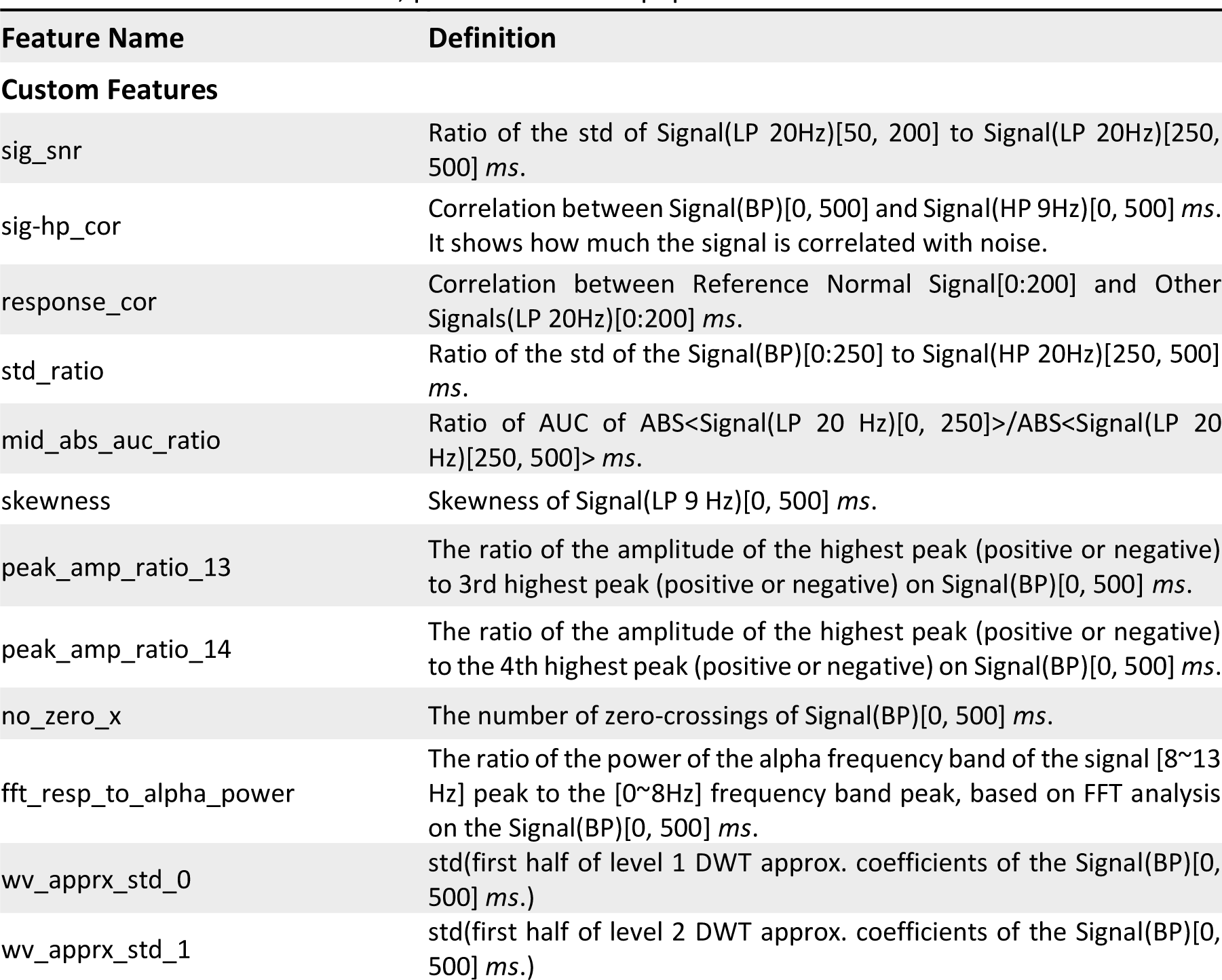
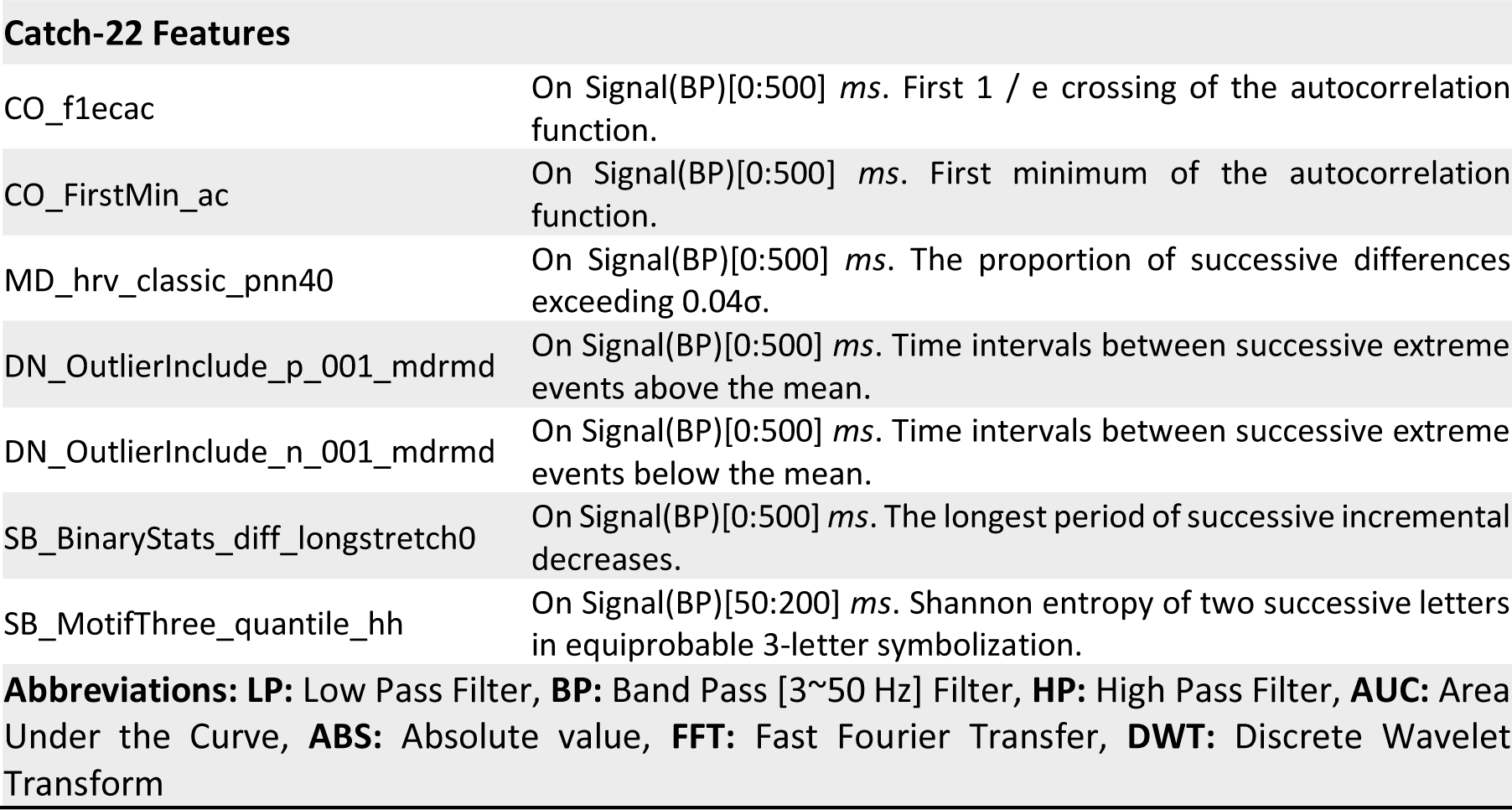
List of features extracted from the signals for the machine learning training. For the details of the Catch-22 features, please refer to its paper.

| Feature Name | Definition |
| --- | --- |
| **Custom Features** |  |
| sig_snr | Ratio of the std of Signal(LP 20Hz)[50, 200] to Signal(LP 20Hz)[250, 500] *ms*. |
| sig-hp_cor | Correlation between Signal(BP)[0, 500] and Signal(HP 9Hz)[0, 500] *ms*. It shows how much the signal is correlated with noise. |
| response_cor | Correlation between Reference Normal Signal[0:200] and Other Signals(LP 20Hz)[0:200] *ms*. |
| std_ratio | Ratio of the std of the Signal(BP)[0:250] to Signal(HP 20Hz)[250, 500] *ms*. |
| mid_abs_auc_ratio | Ratio of AUC of ABS<Signal(LP 20 Hz)[0, 250]>/ABS<Signal(LP 20 Hz)[250, 500]> *ms*. |
| skewness | Skewness of Signal(LP 9 Hz)[0, 500] *ms*. |
| peak_amp_ratio_13 | The ratio of the amplitude of the highest peak (positive or negative) to 3rd highest peak (positive or negative) on Signal(BP)[0, 500] *ms*. |
| peak_amp_ratio_14 | The ratio of the amplitude of the highest peak (positive or negative) to the 4th highest peak (positive or negative) on Signal(BP)[0, 500] *ms*. |
| no_zero_x | The number of zero-crossings of Signal(BP)[0, 500] *ms*. |
| fft_resp_to_alpha_power | The ratio of the power of the alpha frequency band of the signal [8~13 Hz] peak to the [0~8Hz] frequency band peak, based on FFT analysis on the Signal(BP)[0, 500] *ms*. |
| wv_apprx_std_0 | std(first half of level 1 DWT approx. coefficients of the Signal(BP)[0, 500] *ms*.) |
| wv_apprx_std_1 | std(first half of level 2 DWT approx. coefficients of the Signal(BP)[0, 500] *ms*.) |
| Catch-22 Features |  |
| CO_f1ecac | On Signal(BP)[0:500] *ms*. First 1 / $e$ crossing of the autocorrelation function. |
| CO_FirstMin_ac | On Signal(BP)[0:500] *ms*. First minimum of the autocorrelation function. |
| MD_hrv_classic_pnn40 | On Signal(BP)[0:500] *ms*. The proportion of successive differences exceeding $0.04\sigma$. |
| DN_OutlierInclude_p_001_mdrmd | On Signal(BP)[0:500] *ms*. Time intervals between successive extreme events above the mean. |
| DN_OutlierInclude_n_001_mdrmd | On Signal(BP)[0:500] *ms*. Time intervals between successive extreme events below the mean. |
| SB_BinaryStats_diff_longstretch0 | On Signal(BP)[0:500] *ms*. The longest period of successive incremental decreases. |
| SB_MotifThree_quantile_hh | On Signal(BP)[50:200] *ms*. Shannon entropy of two successive letters in equiprobable 3-letter symbolization. |

**Abbreviations:** **LP:** Low Pass Filter, **BP:** Band Pass [3~50 Hz] Filter, **HP:** High Pass Filter, **AUC:** Area Under the Curve, **ABS:** Absolute value, **FFT:** Fast Fourier Transfer, **DWT:** Discrete Wavelet Transform

#### Classification Method

For the classification task, we compared several supervised ML algorithms: Support Vector Machine (SVM), Logistic Regression, K-Nearest Neighbors (KNN), Naïve Bayes, Random Forest, Decision Tree, and ensemble meta-estimators AdaBoost and Bootstrap Aggregating (Bagging) - both of which used Decision Tree as the base estimator. Additionally, we tried a deep learning Neural Networks (NN) algorithm that used an architecture consisting of several stages of one-dimensional convolutional layers terminating with a densely connected classifier network – the highest accuracy that was attained by the NN approach was around 88%. In the end, we selected the SVM binary classifier as it produced the highest accuracy level and a balanced sensitivity vs. specificity. SVM uses hyperplanes to separate the classes with maximal margins. Our model uses the Radial Basis Function (RBF) kernel to conform to the nonlinearities of the classification task.

Most models performed very well on the augmented training data set. For example, a 5-fold cross-validation for the SVM model returns accuracies of [0.9983, 0.9979, 0.9978, 0.9981, 0.9975]. However, we selected our model based on its accuracy on the evaluation dataset. As discussed before, the accuracy of the evaluation set represents not only the accuracy of the model but also the NeuroVEP device, as it takes into account the abnormalities inherent to the mfVEP test and the device. **Table 3** shows the accuracy, sensitivity, specificity, and Area Under the Receiver Operating Characteristic (ROC) Curve (AUC) for the models built using the training set and tested with the evaluation set.

**Table 3.** Performance comparison of different ML models on the evaluation dataset.

| Model | Accuracy | Sensitivity | Specificity | AUC |
| --- | --- | --- | --- | --- |
| SVM | 0.91 | 0.82 | 0.94 | 0.88 |
| Logistic Regression | 0.86 | 0.83 | 0.88 | 0.85 |
| KNN | 0.83 | 0.86 | 0.82 | 0.84 |
| AdaBoost (Decision Tree) | 0.79 | 0.9 | 0.76 | 0.83 |
| Naive Bayes | 0.77 | 0.9 | 0.73 | 0.81 |
| Bagging (Decision Tree) | 0.77 | 0.89 | 0.72 | 0.81 |
| Random Forest | 0.76 | 0.92 | 0.71 | 0.82 |
| Decision Tree | 0.71 | 0.92 | 0.65 | 0.78 |

**Table 4** shows the effect of data augmentation and the addition of Catch-22 features on the test performance. For all these iterations, the SVM model with RBF kernel is used. It can be seen how making the model more complex step by step increased the accuracy from 0.8 to 0.91.

**Table 4.** The effect of data augmentation and Catch-22 features on the performance of the ML model. The symbol ‘✓’ denotes inclusion.

| Model | Data Augmentation | 12 Custom Features | 7 Catch-22 Features | Accuracy | Sensitivity | Specificity | AUC |
| --- | --- | --- | --- | --- | --- | --- | --- |
| SVM | - | ✓ | - | 0.8 | 0.91 | 0.76 | 0.84 |
| SVM | - | ✓ | ✓ | 0.83 | 0.85 | 0.83 | 0.84 |
| SVM | ✓ | ✓ | - | 0.84 | 0.9 | 0.82 | 0.86 |
| SVM | ✓ | ✓ | ✓ | 0.91 | 0.82 | 0.94 | 0.88 |

## Results and Discussion

### Full-Field VEP Test

**Table 5** summarizes all calculated metrics for the study’s subject pool. As discussed, the latency of the P1 is the most robust biomarker for detecting visual pathway damage. Our analysis, primarily looks at the latency and latency differences of the Oz location for each eye, plus the waveform. The waveform is evaluated by amplitude abnormalities. If they were normal, the subject is labeled as normal. If the waveform and latencies of the Oz location were abnormal, we then use the information from the rest of the sensors at O1 and O2 locations to get a better understanding of the deficit. In some instances where waveforms were abnormal, the interhemispheric (IH) or interocular (IO) ratios were marked as NA; for interhemispheric comparisons, the troubling results were reported as being in the left hemisphere (L), right hemisphere (R), or on both sides (B).

**Table 5.** Summary of the results for all subjects.

| Subject | Sex | Age | Condition | L CA Lat | R CA Lat | IO Lat Diff | L Amp | R Amp | IO Amp Ratio | L IH Amp Ratio | L IH Lat Diff | R IH Amp Ratio | R IH Lat Diff |
| --- | --- | --- | --- | --- | --- | --- | --- | --- | --- | --- | --- | --- | --- |
| S 01 | M | 36~40 | Normal | 105 | 102 | 3 | 9.2 | 9.19 | 1 | 1.55 | 20 | 1.01 | 20 |
| S 02 | M | 31~35 | Normal | 107 | 101 | 6 | 7.1 | 7.49 | 1.05 | 1.14 | 10 | 1.01 | 1 |
| S 03 | M | 66~70 | Normal | 104 | 104 | 0 | 2.89 | 3.82 | 1.32 | 1.1 | 2 | 1.17 | 2 |
| S 04 | M | 66~70 | MS/ON | 108 | 113 | 5 | -0.02 | -0.6 | NA (B) | NA (L) | 10 | NA (B) | 1 |
| S 05 | F | 56~60 | Normal | 99 | 90 | 9 | 3.58 | 1.7 | 2.11 | 1.3 | 15 | 1.2 | 2 |
| S 06 | F | 61~65 | Normal | 107 | 102 | 5 | 2.1 | 2.13 | 1.01 | 1.32 | 4 | 1.32 | 0 |
| S 07 | F | 31~35 | Normal | 96 | 89 | 7 | 1.39 | 2.17 | 1.56 | 1.75 | 0 | 2.65 | 8 |
| S 08 | M | 31~35 | Normal | 112 | 111 | 1 | 5.06 | 4.42 | 1.14 | 1.13 | 7 | 1.01 | 11 |
| S 09 | M | 26~30 | Normal | 109 | 113 | 4 | 5.07 | 4.61 | 1.1 | 1 | 6 | 1 | 9 |
| S 10 | F | 46~50 | ON | 108 | 134 | 26 | 2.2 | 2.04 | 1.08 | 1.25 | 10 | 2.66 | 6 |
| S 11 | F | 26~30 | MS | 130 | 106 | 24 | 1.24 | 2.13 | 1.72 | NA (L) | 31 | NA (L) | 17 |
| S 12 | F | 41~45 | Normal | 116 | 118 | 2 | 2.98 | 1.11 | 2.68 | 1.41 | 5 | 1.04 | 7 |
| S 13 | M | 46~50 | Normal | 100 | 86 | 14 | 3.8 | 3.7 | 1.03 | 1.04 | 7 | 1.07 | 0 |
| S 14 | F | 31~35 | MS | 102 | 93 | 9 | 2.32 | 0.17 | 10.05 | 1.23 | 8 | 2.5 (BR) | 9 |
| **S 15** | F | 51~55 | MS | 107 | 129 | 22 | 3.01 | 1.37 | 2.2 | 1.11 | 23 | 1.35 | 15 |
| **S 16** | F | 56~60 | ON | 136 | 136 | 0 | 5.82 | 6.46 | 1.11 | 1.08 | 1 | 1.45 | 1 |
| **S 17** | F | 36~40 | MS/ON | 103 | 95 | 8 | -0.68 | 1.11 | NA | NA (L) | NA (L) | NA (B) | NA (B) |
| **S 18** | M | 61~65 | MS | 108 | 118 | 10 | 2.42 | 1.65 | 1.47 | 1.12 | 5 | 1.09 | 5 |
| **S 19** | M | 61~65 | MS | 98 | 99 | 1 | 1.37 | 1.98 | 1.45 | 9.24 | 6 | 1.71 | 3 |
| **S 20** | F | 36~40 | MS/ON | 123 | 164 | 41 | 1.27 | 1 | 1.27 | 2.62 | 7 | NA (L) | 25 |

L: Left, R: Right, **CA**: Center Array, **Lat**: Latency, **Diff**: Difference, **Amp**: Amplitude, **IO**: Inter-Ocular, **IH**: Inter-Hemisphere
Color Guide: **Normal** **Borderline** **Abnormal**

We compared our device’s ffVEP latency measurements and diagnosis to those of the SOC ffVEP test, being used in practice by our clinical partners at Tufts Medical Center, Boston, Massachusetts. The SOC test was performed using a concentric needle electrode at the Oz location; the technician determines the number of trials, adding more for weaker signals, and then hand-picks the P1 peaks for which the amplitude and latency are reported – the waveforms are also included in the report. Finally, mainly based on the P1 latencies and to some extent the waveforms, a neurologist provides interpretation of the results. In **Table 6**, NeuroVEP’s diagnosis for all subjects is summarized along with the diagnosis using the SOC device.

**Table 6.**
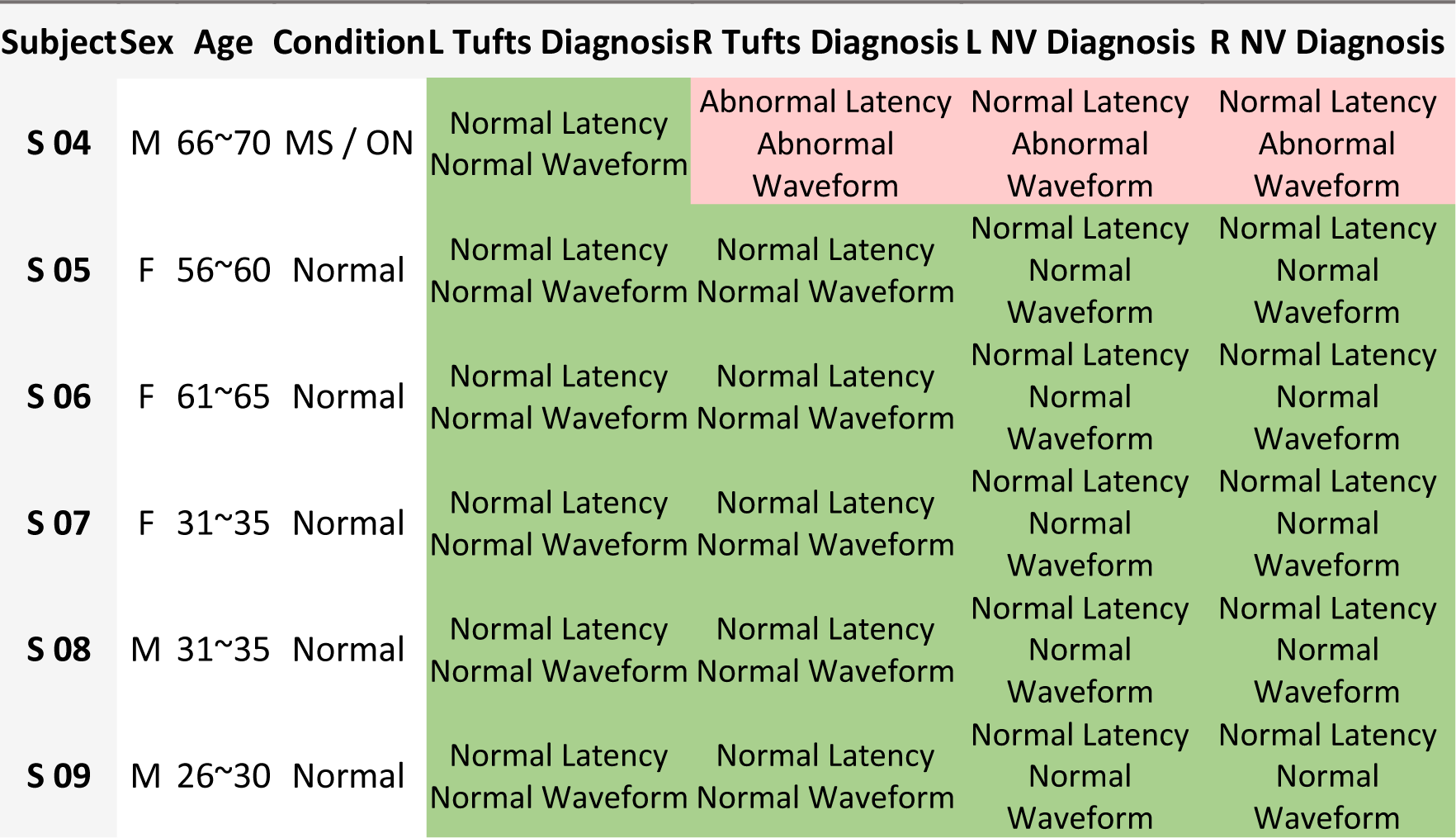
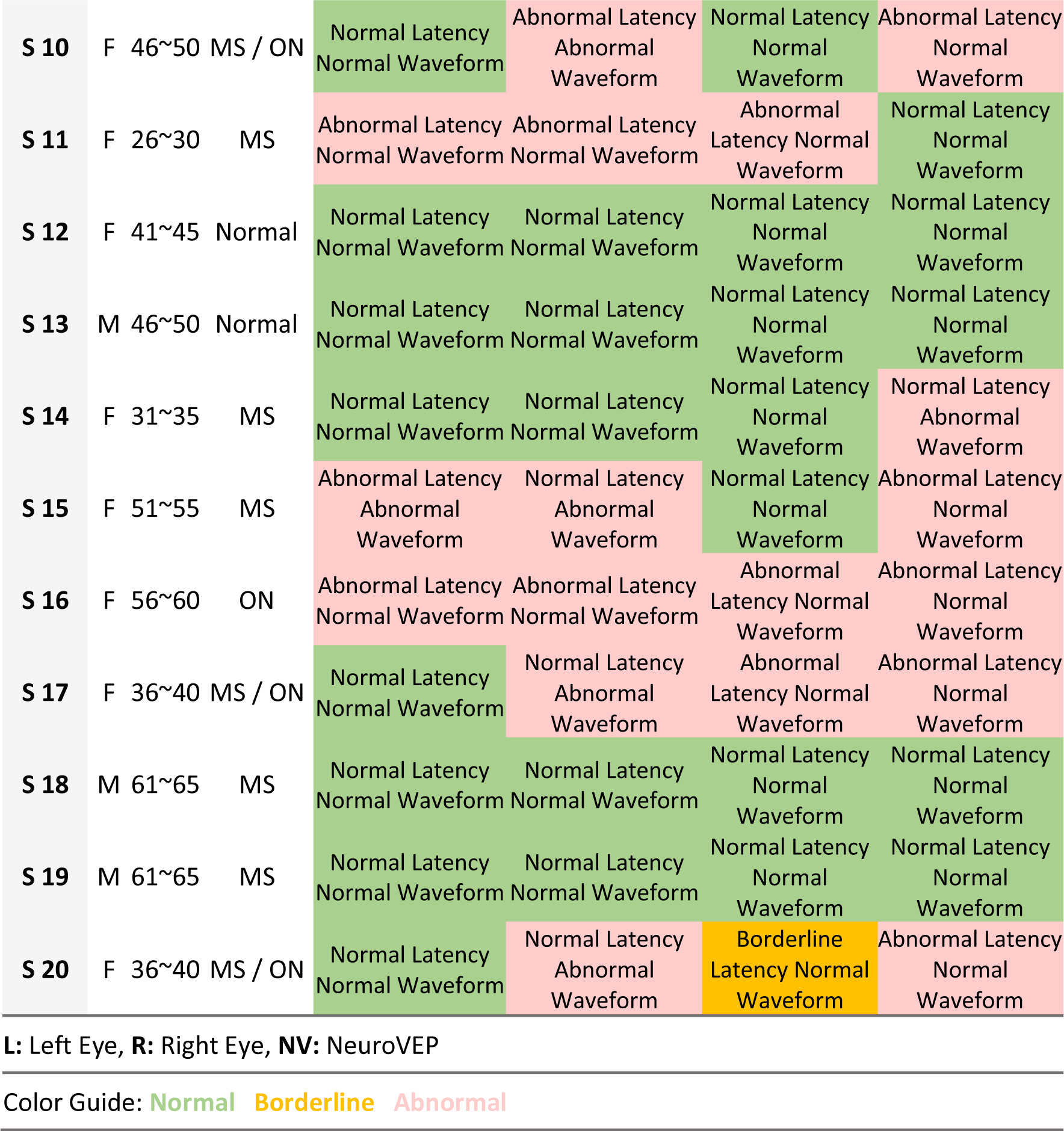
NeuroVEP vs. Tufts Diagnosis Results.

In 31 of 34 eyes (91%), similar predictions were given by NeuroVEP and Tufts Diagnosis. However, the NeuroVEP device provides additional information owing to recording from eight individual sensors. Also, based on the recorded subject’s feedback during the test, we believe NeuroVEP device provides more accurate and reliable measurements. For example, see **Figure 10.A.** (S11/MS), where we can see both responses from the left and right eye on the left hemisphere (O1 Location) have abnormal waveforms (and therefore abnormal interhemispheric amplitude ratios). Because of the additional information here, the issue can be better explained with more detail as a lesion in the chiasmal or post-chiasmal regions of the visual field pathway. In **Figure 10.B.**, the results of the same subject from the SOC device are shown. Here, the technician ran the test 3 times the left eye; all three tests turned out to be successful. For the right eye, the technician ran the test 4 times, but only 3 tests were successful. The latencies are generally variable, and these results are then presented to a neurologist for comments. There are a couple of areas where the NeuroVEP device shines in comparison with the SOC device. The first benefit is convenience, both for the patient and the technician who sets up the device. The NeuroVEP device is a wireless headset that only needs a laptop computer to store the data. The technician just sets up the headset, starts the test, and the rest is done automatically. For the patient, the NeuroVEP device is comfortable and non-invasive. Our EEG electrodes are made of soft hydrogels that are held on the scalp with mild pressure, in contrast to the SOC, which uses needle electrodes inserted into the scalp. Also, comparing **Figure 10.A.** and **10.B.**, we can see other advantages of the NeuroVEP device starting with reliability: our results are based on many more trials which have been filtered with extensive denoising and signal processing methods to get a clear and reliable response. In contrast, the technician runs the SOC test several times with fewer trials and with varying results for the latencies and waveforms. The other key advantage of the NeuroVEP device is the fully automated analysis of the results immediately after the test. All parameters of the signals are validated against our normative dataset and color-coded based on the status of the responses. The NeuroVEP device also provides additional responses from 2 more locations on the scalp (O1 and O2), which can reveal additional insight into the defects on the visual pathway. Also, using multiple electrodes for each site increases the signal quality and reliability of the results.

**Figure 10.**
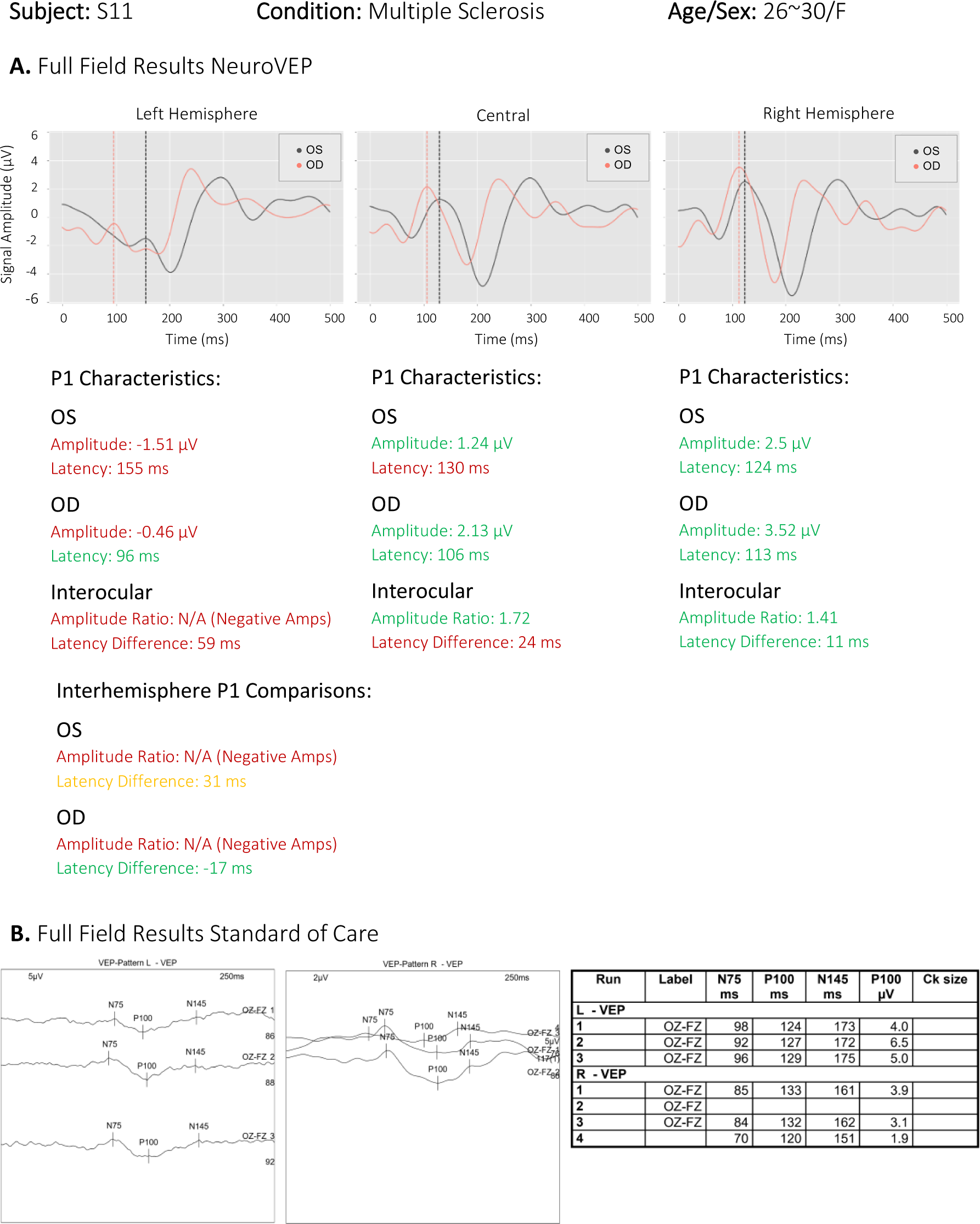
Full-field test reports of a MS/ON subject from **A.** NeuroVEP Device (note, positive peaks point upward) and **B.** Tufts SOC device (note, positive peaks point downward).

### Multi-Focal VEP Test

Interpretation of anomalous mfVEP results is a challenging problem, previously left up to human experts, because signals from poorly sensed (or artificially blocked) sectors often contain residual neural activity, especially intrinsic alpha waves (8-13Hz) and possibly other artifacts which can false-trigger simplistic signal quality metrics. Interocular comparison of the responses has also been proposed as a way to analyze the mfVEP results [36]: since the information about overlapping parts of the visual field from both eyes is analyzed by the same parts of the occipital cortex, the spatial/temporal properties of signals from those collocal cortical regions are often so similar in the case of healthy visual pathways that measured monocular responses often correlate to a high degrees; thus, significant differences in responses between eyes can be valid criteria for identifying damage to the pathways [37]. However, an obvious shortcoming of the interocular method is in cases where deficits exist in both eyes. Incorporating several metrics derived from various unique features of responses while letting an automated ML model classify and score the responses can circumvent some of the shortcomings of simplistic signal metrics and especially improve the monocular analysis of the mfVEP results.

**Figure 11** shows a sample of results for an unaltered stimulus and two different stimuli where defects were simulated (OS: Left eye and OD: Right eye). For each sector, a score between 0 to 100 is calculated based on the machine learning classification probabilities. Scores below 50 are deemed abnormal responses, and above 50 are deemed normal; these scores are used to quantify the sector responses using a grayscale map: whiter sectors are more certainly good responses, darker are more certainly poor responses (or the stimuli were artificially blocked), and mid-gray responses are ambiguous (should be reassessed by a trained technician).

**Figure 11.**
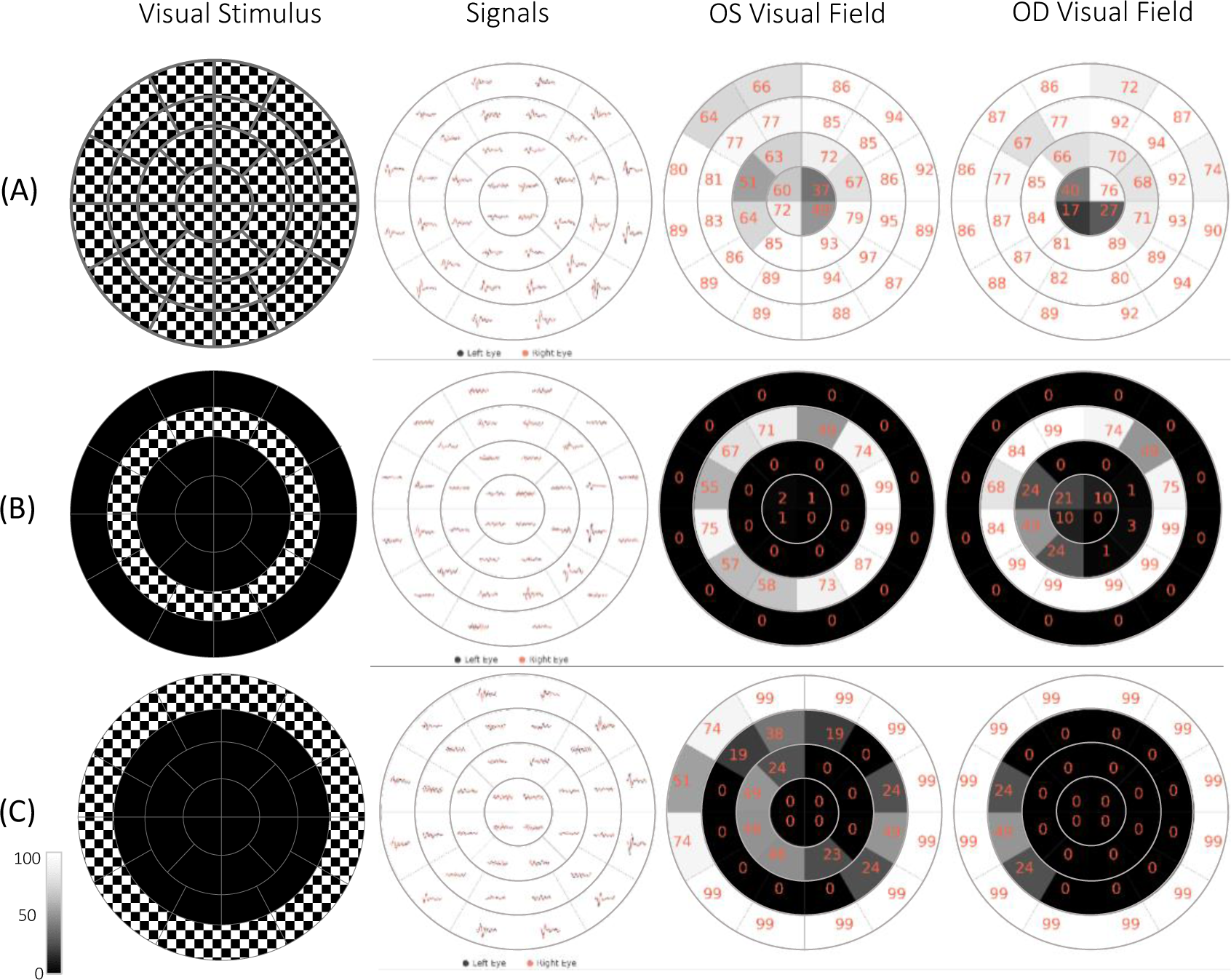
mfVEP custom stimuli, responses, and per eye visual function classifications for **A.** full stimulus 0-22.25° ecc. **B.** Artificially masked mid-peripheral ring stimulus 2.72-8.58° ecc., and **C.** Artificially masked outermost peripheral ring stimulus 8.58-22.25° ecc. Scores between 0 to 100 represent the quality of the response based on the classification probabilities and are linearly gray-scaled (White: Normal, Black: Abnormal); therefore, mid-gray sectors may be marked as “ambiguous”, requiring further analysis.

**Figure 12** shows the ffVEP and mfVEP results of subject S10 with ON. The NeuroVEP’s ffVEP results show abnormal latency for the right eye and an abnormal interhemispheric amplitude ratio. This suggests a dysfunction of the optic nerve on the right side which is probably prechiasmatic. The mfVEP test reveals either very poor vision or complete loss of vision in the dark areas of the right eye’s visual field. The pattern of visual field loss suggests a pre-chiasmatic dysfunction [38]. The NeuroVEP’s diagnosis matches with other clinical tests for this subject. The MRI shows an increased STIR (Short Tau Inversion Recovery) signal abnormality within the right optic nerve and associated enhancements. OCT (Optical Coherence Tomography) test for this subject reveals normal left eye but moderately severe thinning (except nasally) in the right eye.

**Figure 12.**
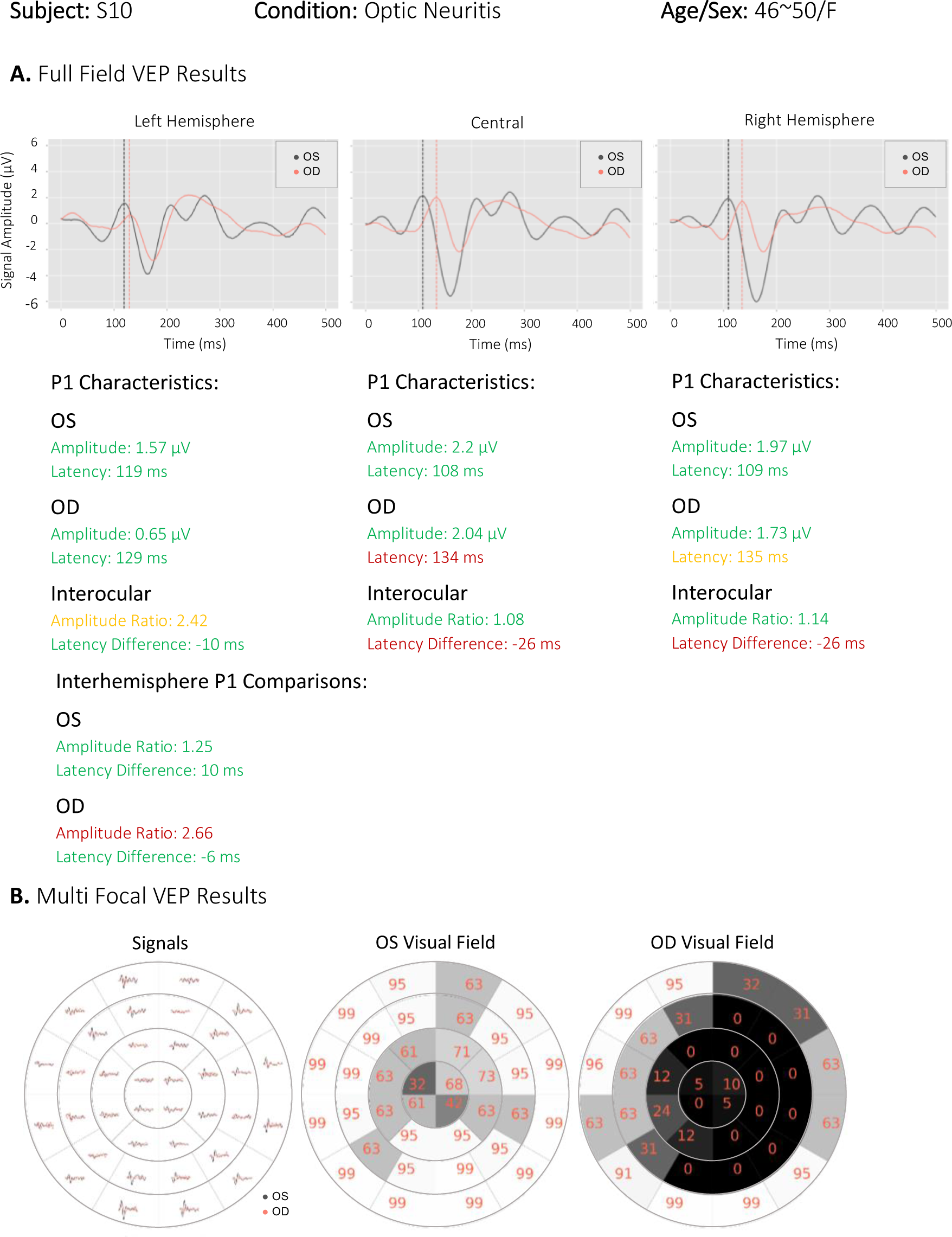
NeuroVEP’s results for subject S10 with right eye (OD) optic neuritis. **A.** ffVEP results with delayed right eye (OD) and abnormal interhemispheric amplitude ratio, **B.** mfVEP results which show a scotoma in the right eye.

**Figure 13** shows the comparison of the NeuroVEP’s mfVEP test results and the HVF (Humphrey Visual Field) test of subject S17. The HVF test is conducted with 30-2 standard, but we have overlayed the location of the 36-sector mfVEP stimulus (which better matches with HVF 24-2 standard test area) on the HVF results to be able to compare the two tests. Optic neuritis in the right eye (OD) of this subject was confirmed by MRI. The optometrist reports mention vision loss in OD and HVF confirms a cecocentral scotoma. The subject can distinguish colors in the right eye but cannot see shapes. This subject expressed extreme discomfort with the HVF test, but was very satisfied with the easiness and objectivity of the NeuroVEP’s visual field test.

**Figure 13.**
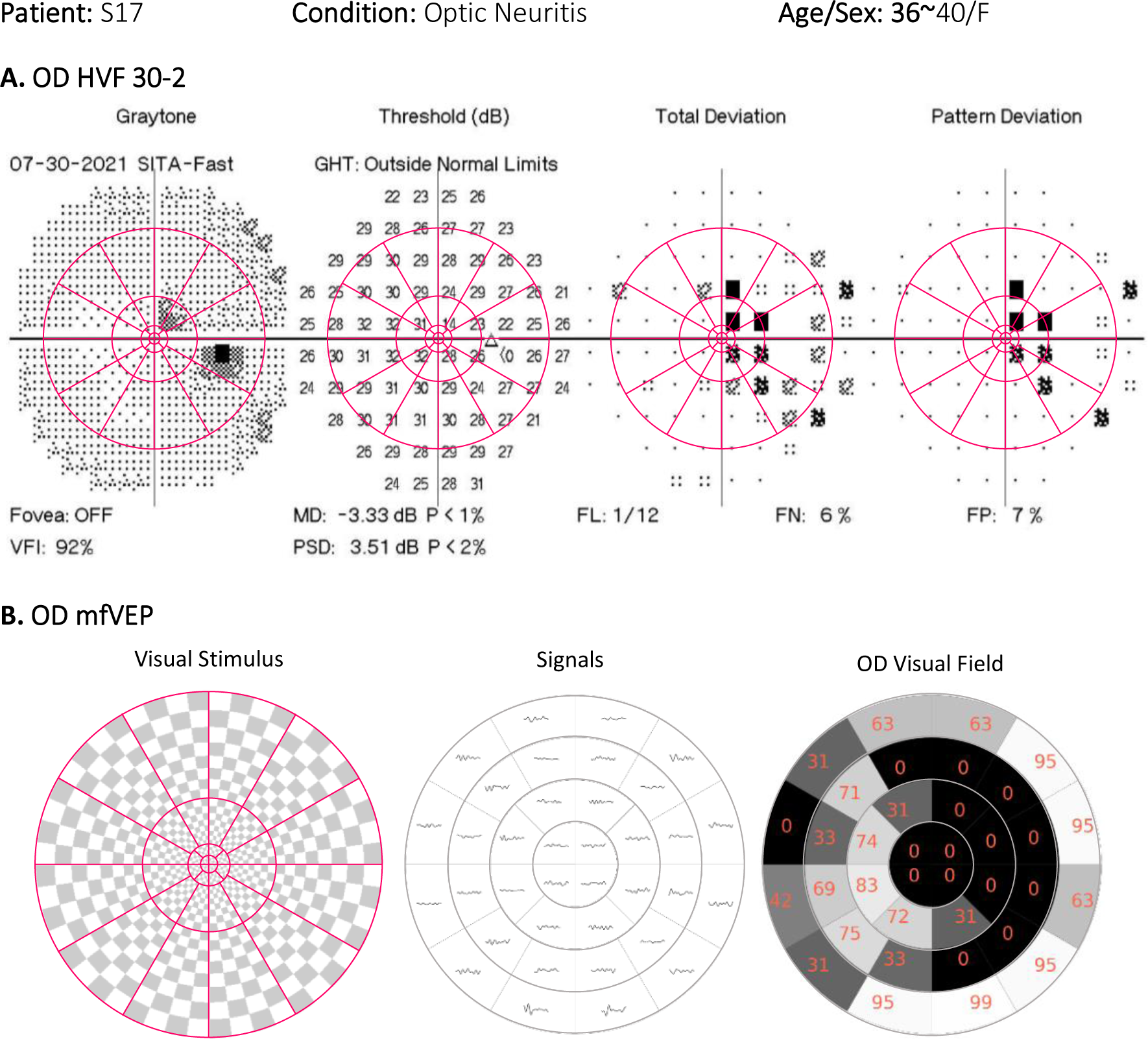
**A.** HVF Central 30-2 Threshold Test with mfVEP sectors’ locations overlayed on top, **B.** NeuroVEP mfVEP test for right eye (OD) of an ON subject. Brain MRI Results: Optic Neuritis Optometrist Notes: Vision loss in right eye (OD), can see some colors, not shapes. HVF: Cecocentral Scotoma. NeuroVEP ffVEP: Abnormal waveform for the right eye (OD). Probably a prechiasmatic dysfunction. NeuroVEP mfVEP: Very poor or loss of vision in the black area.

Our method for classifying the mfVEP responses has its limitations too. One of the limitations is that the two outermost peripheral rings produce much higher SNR responses than the central sectors; this discrepancy was also observed by other researchers [6]; lumping all sectors together for classification, as we have done here, and also the small number of original samples may have introduced some bias that skews more accurate in the periphery. To sum up, by introducing more example responses per sector and tweaking our ML training approach, we believe that ambiguity and false negatives can be reduced even further.

## Conclusion

Diseases of the afferent visual system are central to the practice of neuro-ophthalmology (N-O). Right now, clinicians must perform screening using automated perimetry (AP) and optical coherence tomography (OCT) on patients every 3-6 months typically for patients with such neuro-ophthalmic diseases. AP is subjective, susceptible to various artifacts, has poor accuracy and reliability, and does not provide crucial discriminatory information on the function of the neuro-optical pathways through the brain. OCT provides only structural information about the retina and optic disc (the proximal start of the optic nerve fibers). Visual function often dictates the type of treatment, but screening exams are a burden on already stretched neuro-ophthalmology clinics. Therefore, there is an unmet need for a practical, rapid, sensitive test that is objective and provides quantitative functional endpoints for early diagnosis and for monitoring patient response to newly developed treatments for neuro-ophthalmic diseases.

We have developed a new neuro-ophthalmic testing system called NeuroVEP that combines high-resolution sensors for scalp electric potentials and fields with precise visual stimuli in a portable, mobile computing enabled form-factor. NeuroVEP noninvasively measures VEP over the subject’s visual cortex, sensing its response to custom visual stimulus patterns presented on a head-mounted smartphone display. The neuroelectric recordings have been analyzed for full-field stimulus (ffVEP) and 36 visual field sectors (mfVEP). The ff/mfVEP test is monocular, typically taking about 15 minutes to examine both eyes, and is completely objective, requiring no behavioral response from the subject – just fixation. Our study of ON / MS patients demonstrated the superiority of the NeuroVEP system over a conventional wired clinical VEP system in terms of portability and ease of use, superior VEP results, in addition to providing mfVEP visual field results. The mfVEP results were evaluated using artificial defects introduced to the visual stimulus and our machine learning algorithm achieved 91% classification accuracy for mfVEP sectors.

We validated the NeuroVEP’s system in a study of 20 subjects (10 healthy, 10 MS and/ or ON patients). NeuroVEP’s ffVEP results were compared with the performance of the SOC VEP testing device at our clinical partners’ clinic and the diagnosis reported by both systems agreed in 91% of cases. Where available, the NeuroVEP mfVEP results were in good agreement with Humphrey Automated Perimetry visual field analysis and with MRI and OCT findings. This pilot study indicates that NeuroVEP has the potential to be a reliable, portable, and objective diagnostic device for electrophysiology and visual field analysis for neuro-visual disorders.

Despite high accuracies achieved in both ffVEP and mfVEP tests, certain modifications can further improve the reliability, performance, and accuracy of the NeuroVEP device. For ffVEP and especially mfVEP tests, fixation is very important. Like any other visual field test, loss of fixation is detrimental to the test’s result. Therefore, an active eye tracker which tracks the gaze location during stimulation trials is a crucial add-on to the headset. Currently, we use only the live video feed from the Pupil Labs eye-tracking cameras inside the headset, but the test administrator is responsible for making sure the subject is compliant with the test’s protocols. Unfortunately, we found that this 3^rd^ party eye-tracking device was cumbersome to set up and the data was often too noisy to be used for our purposes. A well-integrated eye-tracking solution that tracks the pupil direction, not only can eliminate the need for constant attention of the test administrator but also can provide a reliability metric for the test. Developing a reliability metric for the test, either based on the noise level of signals, alpha wave contamination, pupil location, or a fusion of all these parameters, is an important step in improving the NeuroVEP headset as a diagnostic apparatus. Increasing the number of stimulating sectors to 60 or more can increase the test’s resolution. Another promising area of improvement is the machine learning classification training procedure; as discussed in the previous sections, we lumped together signals from various eccentricities. However, we do not get the same quality of signals from all eccentricities. Peripheral ring sectors tend to produce higher SNR responses, and the central sectors tend to produce lower ones. Classifying all sectors using the same criteria can create a bias towards better responses in the periphery. Since we eventually use the classification probabilities for scoring the sector responses, a threshold moving algorithm can improve the slightly imbalanced classification problem. However, we believe obtaining more samples by testing more subjects will enable us to break up the classification task into several sub-tasks using each set of sectors within the same eccentricity ring separately; this strategy may reduce biases and improve the overall accuracy. Furthermore, although interocular comparisons of the responses may have shortcomings if used as the singular criterion for the classification, being able to incorporate them in the ML framework should noticeably increase the accuracy of the classifications.

## Data Availability

All data produced in the present work are contained in the manuscript.

## Acknowledgements

This work was partially supported by the National Institutes of Health CTSI grant UL1TR002544. We would like to thank Dr. Srinivasan Radhakrishnan for fruitful discussions on the data analysis of mfVEP.

## REFERENCES

1. Barton, J.L., et al., The electrophysiological assessment of visual function in Multiple Sclerosis. Clinical neurophysiology practice, 2019. 4: p. 90–96.

2. Hartung, D.M., Economics and cost-effectiveness of multiple sclerosis therapies in the USA. Neurotherapeutics, 2017. 14(4): p. 1018–1026.

3. Halliday, A., W. McDonald, and J. Mushin, Delayed visual evoked response in optic neuritis. The Lancet, 1972. 299(7758): p. 982–985.

4. Baseler, H., et al., The topography of visual evoked response properties across the visual field. Electroencephalography and clinical Neurophysiology, 1994. 90(1): p. 65–81.

5. Hood, D.C., J.G. Odel, and X. Zhang, Tracking the recovery of local optic nerve function after optic neuritis: a multifocal VEP study. Investigative ophthalmology & visual science, 2000. 41(12): p. 4032–4038.

6. Hood, D.C. and V.C. Greenstein, Multifocal VEP and ganglion cell damage: applications and limitations for the study of glaucoma. Progress in retinal and eye research, 2003. 22(2): p. 201–251.

7. Betsuin, Y., et al., Clinical application of the multifocal VEPs. Current eye research, 2001. 22(1): p. 54–63.

8. Buitinck, L., et al., API design for machine learning software: experiences from the scikit-learn project. arXiv preprint arXiv:1309.0238, 2013.

9. Virtanen, P., et al., SciPy 1.0: fundamental algorithms for scientific computing in Python. Nature methods, 2020. 17(3): p. 261–272.

10. Versek, C., et al., Portable diagnostic system for age-related macular degeneration screening using visual evoked potentials. Eye and brain, 2021. 13: p. 111.

11. Banijamali, S.M.A., Portable Brain and Vision Diagnostic System for Age-Related Macular Degeneration and Multiple Sclerosis/Optic Neuritis. 2023, Northeastern University: United States -- Massachusetts. p. 126.

12. Kassner, M., W. Patera, and A. Bulling. Pupil: an open source platform for pervasive eye tracking and mobile gaze-based interaction. in Proceedings of the 2014 ACM international joint conference on pervasive and ubiquitous computing: Adjunct publication. 2014.

13. Versek, C., et al., Electric field encephalography for brain activity monitoring. Journal of neural engineering, 2018. 15(4): p. 046027.

14. Hood, D.C., et al., Visual field defects and multifocal visual evoked potentials: evidence of a linear relationship. Archives of Ophthalmology, 2002. 120(12): p. 1672–1681.

15. Sutter, E.E., Imaging visual function with the multifocal m-sequence technique. Vision research, 2001. 41(10-11): p. 1241–1255.

16. Greenfield, P., M. Droettboom, and E. Bray, ASDF: A new data format for astronomy. Astronomy and computing, 2015. 12: p. 240–251.

17. Odom, J.V., et al., Visual evoked potentials standard (2004). Documenta ophthalmologica, 2004. 108: p. 115–123.

18. Pedregosa, F., et al., Scikit-learn: Machine learning in Python. the Journal of machine Learning research, 2011. 12: p. 2825–2830.

19. Aminoff, M.J., Aminoff’s Electrodiagnosis in Clinical Neurology: Expert Consult-Online and Print. 2012: Elsevier Health Sciences.

20. Berger, H., Über das elektrenkephalogramm des menschen. DMW-Deutsche Medizinische Wochenschrift, 1934. 60(51): p. 1947–1949.

21. Kropotov, J., Quantitative EEG, event-related potentials and neurotherapy. 2010: Academic Press.

22. Halliday, A., W. McDonald, and J. Mushin, Visual evoked response in diagnosis of multiple sclerosis. Br Med J, 1973. 4(5893): p. 661–664.

23. Ghilardi, M.F., et al., N70 and P100 can be independently affected in multiple sclerosis. Electroencephalography and Clinical Neurophysiology/Evoked Potentials Section, 1991. 80(1): p. 1–7.

24. Weinstock-Guttman, B., et al., Pattern reversal visual evoked potentials as a measure of visual pathway pathology in multiple sclerosis. Multiple Sclerosis Journal, 2003. 9(5): p. 529–534.

25. Society, A.C.N., Guideline 9B: guidelines on visual evoked potentials. American journal of electroneurodiagnostic technology, 2006. 46(3): p. 254–274.

26. Sutter, E.E., The fast m-transform: a fast computation of cross-correlations with binary m-sequences. SIAM Journal on Computing, 1991. 20(4): p. 686–694.

27. Klistorner, A. and S.L. Graham, Objective perimetry in glaucoma. Ophthalmology, 2000. 107(12): p. 2283–2299.

28. Steinmetz, H., G. Fürst, and B.-U. Meyer, Craniocerebral topography within the international 10– 20 system. Electroencephalography and clinical neurophysiology, 1989. 72(6): p. 499–506.

29. Hood, D.C., et al., Quantifying the benefits of additional channels of multifocal VEP recording. Documenta Ophthalmologica, 2002. 104(3): p. 303–320.

30. Meigen, T. and M. Krämer, Optimizing electrode positions and analysis strategies for multifocal VEP recordings by ROC analysis. Vision research, 2007. 47(11): p. 1445–1454.

31. Zhang, X., et al., A signal-to-noise analysis of multifocal VEP responses: an objective definition for poor records. Documenta Ophthalmologica, 2002. 104(3): p. 287–302.

32. Maneewongvatana, S. and D.M. Mount, Analysis of approximate nearest neighbor searching with clustered point sets. arXiv preprint cs/9901013, 1999.

33. Goldberg, I., S.L. Graham, and A.I. Klistorner, Multifocal objective perimetry in the detection of glaucomatous field loss. American Journal of Ophthalmology, 2002. 133(1): p. 29–39.

34. Hood, D.C., X. Zhang, and B.J. Winn, Detecting glaucomatous damage with multifocal visual evoked potentials: how can a monocular test work? Journal of Glaucoma, 2003. 12(1): p. 3–15.

35. Lubba, C.H., et al., catch22: CAnonical Time-series CHaracteristics: Selected through highly comparative time-series analysis. Data Mining and Knowledge Discovery, 2019. 33(6): p. 1821–1852.

36. Hood, D.C., et al., An interocular comparison of the multifocal VEP: a possible technique for detecting local damage to the optic nerve. Investigative ophthalmology & visual science, 2000. 41(6): p. 1580–1587.

37. Remington, L.A. and D. Goodwin, Clinical Anatomy and Physiology of the Visual System E-Book. 2021: Elsevier Health Sciences.

38. Ettinger, A.B. and D.M. Weisbrot, Neurologic differential diagnosis: a case-based approach. 2014: Cambridge University Press.

